# Surveillance of potential pathogens and antibiotic resistance in wastewater and surface water from Boston, USA and Vellore, India using long-read metagenomic sequencing

**DOI:** 10.1101/2021.04.22.21255864

**Authors:** Erica R. Fuhrmeister, Lee E. Voth-Gaeddert, Angeline Metilda, Albert Tai, Rebecca E. Batorsky, Balaji Veeraraghavan, Honorine D. Ward, Gagandeep Kang, Amy J. Pickering

## Abstract

Environmental sampling (wastewater) could be an efficient surveillance strategy to capture global emerging trends in the spread of antibiotic resistance. Long-read DNA sequencing can resolve the genetic context of antibiotic resistance genes (ARGs) and is a promising tool for non-culture-based monitoring of antibiotic-resistant pathogens and ARGs in environmental samples, but has not been rigorously validated against conventional methods. We tested long-read sequencing using the portable Nanopore MinION for surveying pathogens, ARGs, and antibiotic-resistant pathogens in municipal wastewater, hospital wastewater, and surface water collected from Boston, USA and Vellore, India. We compared detection of enteric pathogens by assembly of long reads, with and without short-read polishing, and unassembled raw long reads for ARGs to multiplex real-time PCR. Using real-time PCR as a benchmark, long-read metagenomics was 49% sensitive and 75% specific at pathogen detection in assembled contigs, and 16% sensitive and 100% specific at detecting 28 clinically relevant resistance genes in raw long reads. Short-read polishing did not substantially improve pathogen identification or impact ARG identification in the assembled contigs, demonstrating that short-read polishing is not required, which greatly reduces costs. The high specificity of ARG detection supports portable long-read sequencing as a valuable tool to profile ARGs and antibiotic-resistant pathogens for environmental surveillance programs.

## Introduction

Globally, the proliferation of antibiotic resistance remains a critical threat to public health. Antibiotic-resistant infections are more prone to treatment failure, which in turn increases mortality rates, treatment time, and health care costs^1^. Antibiotic-resistant bacteria (ARB) have been documented in the environment (e.g. soils, surface water, sediments), particularly in locations with anthropogenic pressures such as antibiotic residues in wastewater outflows^2, 3^. However, human and environmental reservoirs of ARB are not well understood in part because there has been limited standardized monitoring. Existing antibiotic surveillance systems are largely limited to isolates from clinical settings^4, 5^. Antibiotic resistance compounds the threat of enteric pathogen infection (e.g. *Shigella*, *Salmonella typhi*, *Helicobacter pylori*) which is common in many low- and middle-income countries^6–8^. Similar to antibiotic resistance, there are few standardized monitoring systems in place for enteric pathogens in high- or low-income countries. The WHO Global Foodborne Infections Network^9^ is a promising surveillance system that surveys isolates of antibiotic-resistant enteric pathogens in humans, animals, and the environment but has been limited to selected pathogens.

The rapid global expansion of wastewater surveillance for monitoring SARS-CoV-2^10^ presents a unique opportunity to implement a system for antibiotic resistance and other pathogens. Surveying wastewater is an efficient way to monitor enteric pathogens and antimicrobial resistance excreted in feces over large populations^11^. A robust surveillance system would also include environmental reservoirs (e.g. soil and water) as well as animals to encompass a “One Health” approach^12^. One barrier to implementing monitoring systems is selecting appropriate methods for detection. Previous studies have used diverse methods to detect antibiotic resistance genes (ARG) in wastewater, sludge, soil, water, and sediment, making it difficult to compare results across locations and sample types^13^. Culture-based methods are able to identify phenotypic resistance, however most bacteria are not easily cultured and the number of isolates examined can vary widely across studies ^13^. Polymerase chain reaction (PCR) can detect low abundance genes, but targets must be pre-specified prior to analysis and determining co-occurrence of multiple genes within the same organism (e.g. pathogen gene and antibiotic resistance gene located in the same genome) is not possible. High-throughput sequencing is a promising tool for building improved antibiotic resistance monitoring systems ^14^. Gene targets do not need to be pre-specified which allows for broad assessments of ARG diversity. However, for the most common platforms (e.g. Illumina), startup costs remain high, turnout time can be slow, devices are not easily moveable, and short DNA reads are limited in their ability to reveal the genetic context of ARGs^15, 16^.

The Oxford Nanopore Technologies (ONT) MinION is a pocket-sized, portable sequencing device that has been used for real-time detection of pathogens during outbreaks (e.g. Zika virus, SARS-Cov-2)^17, 18^ and to detect ARGs in cultured isolates^19^. Nanopore sequencing differs from other next-generation technologies in that DNA is run through protein pores, changing the electrical current which is analyzed to determine the sequence^20^. The small size and portability of the device renders it a good candidate for surveillance applications around the world. While there are significant costs associated with flow-cells and reagents, there is no large capital investment needed for the sequencing devices as opposed to other metagenomic sequencing technologies (Illumina). However, there has been limited application of the Nanopore MinION for detection of pathogens or ARGs from environmental water samples and few studies have used metagenomic long-read sequencing (as opposed to amplicon long-read sequencing) ^21–24^.

In the study of antibiotic resistance genes in the environment, there is a need to link resistance genes to their host organisms in order to identify pathogens carrying resistance and evaluate their genetic context. While long reads are promising in this area, the read length obtained can vary significantly depending on DNA extraction procedures and library preparation protocols. For example, bead beating, a common procedure used to lyse DNA in hardy environmental samples (e.g. wastewater, activated sludge, soil), can shear DNA fragments. We therefore investigated the utility of assembled long reads for surveillance since assembly can improve taxonomic classification and identification of mobile genetic elements (plasmids)^25^. In this study, we compare long-read sequencing via the Nanopore MinION to real-time PCR for identifying pathogens and ARGs in environmental waters. We also compare identification of enteric pathogens and pathogens from the CDC threat list for ARGs with and without short-read polishing of long-read assembled contigs. Furthermore, we test these methods across different water sample types including wastewater, surface water, and hospital wastewater from Boston, USA and Vellore, India. In India, several recent studies reported high levels of ARGs in child stool samples and in wastewater^26, 27^. Prevalence and diversity of resistance genes varies geographically^14^, and the ARGs and antibiotic-resistant bacteria causing resistant infections in the US may be different than those in India^28^.

## Materials and Methods

### Sampling Sites

Environmental samples were collected from the greater Boston area, in Massachusetts, USA (n=6; October 2017) and within the city of Vellore, in Tamil Nadu, India (n=6; April 2018). The sampling locations included three different types of environmental samples hypothesized to have different profiles of ARGs and microbial taxa. Boston sampling included wastewater (influent) entering three wastewater treatment plants (WWTPs), two surface water samples from rivers (downstream of the WWTP discharge points), and one hospital wastewater sample (collected before entering the combined public sewer). Vellore sampling included wastewater (influent) entering two WWTPs, three surface water samples from drainage canals, and one hospital wastewater sample (collected before onsite treatment occurred).

### Sample Collection, Processing, and DNA Extraction

Whirl Pak bags (Nasco, Modesto, CA) were used to collect 2.5L of water from each sampling site. Samples were concentrated by 0.22 µm Nanoceram (Argonide, Sanford, FL) disc filters and DNA was extracted from filters using the Qiagen DNeasy PowerWater kit (Germantown, MD). A mock mixed culture of six bacterial species with and without antibiotic resistance was developed by the Christian Medical College (CMC) laboratory for analysis by long-read sequencing (Table S1). Further details on sample collection, processing, and DNA extraction can be found in the supplementary material.

### Molecular Analysis

DNA extract for each sample was analyzed using the TaqMan Array Card (TAC)^29^ which includes 19 bacterial pathogen gene targets; 39 targets total, excluding RNA viruses (Table S2). Samples were also analyzed using a real-time PCR antibiotic resistance array card (Qiagen, Cat. no. 330261 BAID-1901ZRA), which has 83 ARG targets (Table S3). Long-read sequences were obtained using an R9.4.1 flow cell on Oxford Nanopore Technology’s (ONT) MinION (Oxford, United Kingdom). Library preparation was conducted using the SQK-LSK109 ligation sequencing kit (ONT) according to the manufacturer’s instructions. DNA extracts of all samples, except for the mock community, were sequenced on an Illumina HiSeq2500 (PE150 Rapid V2) by the Tufts Genomic Core Facility. The Nextera DNA XT kit (Illumina, San Diego, CA) was used for library preparation. The datasets generated in this work are available in NCBI’s Sequence Read Archive under BioProject PRJNA672704.

### Data Analysis

Long read sequences were basecalled and trimmed using Guppy (ONT, v3.3.0). Any reads with a quality score < 7 were removed^30^. Bbmap (v38.7)^31^ was used to remove adapters and PhiX sequences from Illumina short reads and sickle^32^ was used for quality trimming. Long read sequences were assembled using metaFlye (v2.6) ^33^ and polished using Racon (v1.4.13)^34^, followed by Medaka (ONT, v0.11.0). For short-read polishing, contigs were polished again with Racon using Illumina reads. In this study, “polished contigs” or “short-read polishing” refers to assembled contigs that underwent the additional polishing step, where short reads are aligned to the long-read assembly in order to improve the sequence accuracy, whereas “unpolished contigs” refers to contigs that did not undergo polishing with short reads.

ARGs were identified by aligning assembled contigs to the ResFinder database^35^ (duplicates removed) using Minimap2 (v2.17-r941) ^36^. Only primary ARG alignments with ≥90 % identity and ≥100 bp alignment lengths were retained for analysis. To identify taxonomic classification, assembled contigs were aligned to the National Center for Biotechnology Information’s (NCBI’s) RefSeq database^37^ (plasmid sequences removed) using Centrifuge (v1.0.4)^38^. MOB-suite (v1.4.9) was used to identify contigs as plasmids^25^.

To calculate abundance of resistance genes and compare ARG identification between long-read sequencing and real-time PCR, long reads, without short-read polishing, were aligned to the ResFinder database as described above. For absolute abundance, ARG counts were normalized by gigabase pairs (Gbp) of data classified as bacterial taxa when aligned to the RefSeq database as described above. All data analysis was conducted in R (v3.5.0). In our analysis of potential pathogens, we included contigs that were classified as *A. hydrophila*, *B. fragilis*, *C. jejuni*, *C. coli*, *C. difficile*, *H. pylori*, *Salmonella* spp., *V. cholerae*, *A. baumannii*, *E. feacium*, *K. pneumoniae*, *Shigella* spp., *S. aureus*, *S. pneumoniae*, and *P. aeruginosa*. This set of pathogens encompasses bacterial pathogen targets included on the pathogen array card and those on the CDC threat list for antibiotic resistance^39^. In our analysis comparing long-read sequencing to real-time PCR for ARG identification, we selected a subset 28 resistance genes from the array card to compare to Resfinder based on known clinical relevance.

In order to compare detection of pathogens by real-time PCR and long-read sequencing, we calculated sensitivity and specificity using real-time PCR as the gold standard. Here, sensitivity was defined as the true positive rate and specificity as the true negative rate. To evaluate pathogen classification we used the Centrifuge score, or sum of squared lengths of matched segments^38^. Significant differences between unpolished and polished contigs were assessed using log10 transformed Centrifuge scores and paired t-tests. When comparing scores for individual pathogens, p-values were adjusted to correct for multiple comparisons (15 pathogens) using the Bonferroni method^40^. To compare antibiotic resistance identification between unpolished and polished contigs, we calculated concordance by dividing the number of times the same ARG was identified in the polished and unpolished contigs by the total number of ARG identifications in the contigs (total hits).

## Results

### Quality Control

An average of 6.6 Gbp were obtained per sample from Illumina sequencing (Table S4). Long-read sequencing yielded an average of 3.4 Gbp of data per sample, excluding the mock community. All real-time PCR NTCs and extraction blanks were negative for pathogen gene targets. The resistance gene *ermB* amplified in the real-time PCR no template control and was therefore excluded from the analysis. All contigs assembled from long reads in the mock community sample containing ARGs were identified as one of the five expected taxa at the species or subspecies level or as a plasmid (Figure S1). Of the 75 contigs assembled in the mock community sample, including contigs without ARGs, 72 were classified as one of the expected six taxa (five with ARGs, one without) at the species or subspecies level.

### Identification of Pathogens

By real-time PCR, Boston samples were positive for, on average, 12 ± 6 out of the 24 possible bacterial pathogen gene targets whereas Vellore samples were positive for 19 ± 1 bacterial pathogen genes (Table S5). Using real-time PCR as a benchmark, long-read metagenomic sequencing was 49% [stdev: 29%] sensitive and 75% [21%] specific at detecting pathogens using polished contigs (Figure 1, and Table S6). A similar sensitivity (50% [24%]) and specificity (79% [16%]) were obtained when using unpolished contigs. Across all samples, specificity and sensitivity varied by organism, with the highest sensitivity in *Aeromonas*, *B. fragilis*, *C. jejuni*/*coli, and C. difficile*. Long read metagenomics identified contigs classified as *Campylobacter jejuni*/*coli*, *Salmonella* species, *H. pylori*, *C. difficile*, and *S. aureus* that were not identified by PCR. Many of these contigs were relatively short (<2000 bps) and had low Centrifuge scores (sum of squared lengths of matched segments^38^); therefore, they are likely false positives (Table S7).

**Figure 1:**
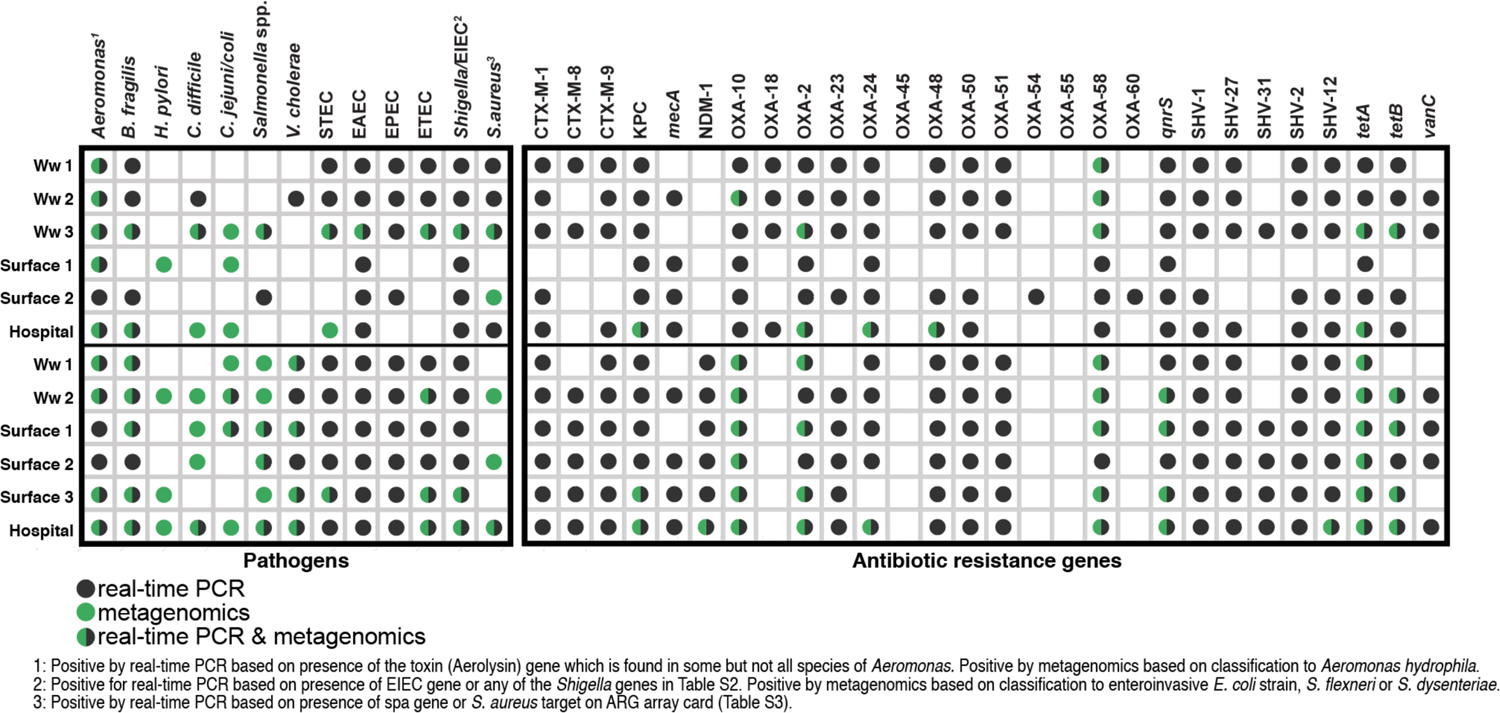
Identification by real-time PCR only is indicated with a gray circle, metagenomics only with a green circle, and both with a green and gray circle. **LEFT**) Pathogens identified using real-time PCR (pathogen genes) and metagenomics in municpal wastewater(ww), hospital wastewater (hospital), and surface water (surface) in Boston and Vellore. Presence via metagenomics was determined by Centrifuge taxonomic classification to the species or strain level (for pathogenic *E. coli*) of contigs assembled from long-reads with short-read polishing. **RIGHT)** Antibiotic resistance genes identified using real-time PCR and long-read sequencing. Samples were considered positive by metagenomics if raw long reads (unpolished) aligned to the equivalent resistance gene in the ResFinder database at an identity > 90% and match length > 100 bp.

The average log10 Centrifuge score was not significantly different between polished and unpolished contigs identified as pathogens (mean polished= 6.3, mean unpolished= 6.0, p-value = 0.08, Table S7). However, when evaluating each pathogen individually, Centrifuge scores were significantly higher for polished versus unpolished paired contigs classified as *A. baumannii*, *A. hydrophila*, *E. coli*, *K. pneumoniae, and P. aeruginosa* (Figure 2; paired t-test, Bonferroni corrected p-value < 0.0001 for all). When including unpaired contigs that changed taxa classification after polishing (Figure S2), there was a significant difference between the unpolished and polished contigs classified as *E. coli* and *K. pneumoniae* (unpaired t-tests, *E. coli* Bonferroni corrected p-value <0.001 and *K. pneumoniae* Bonferroni corrected p-value =0.02).

**Figure 2:**
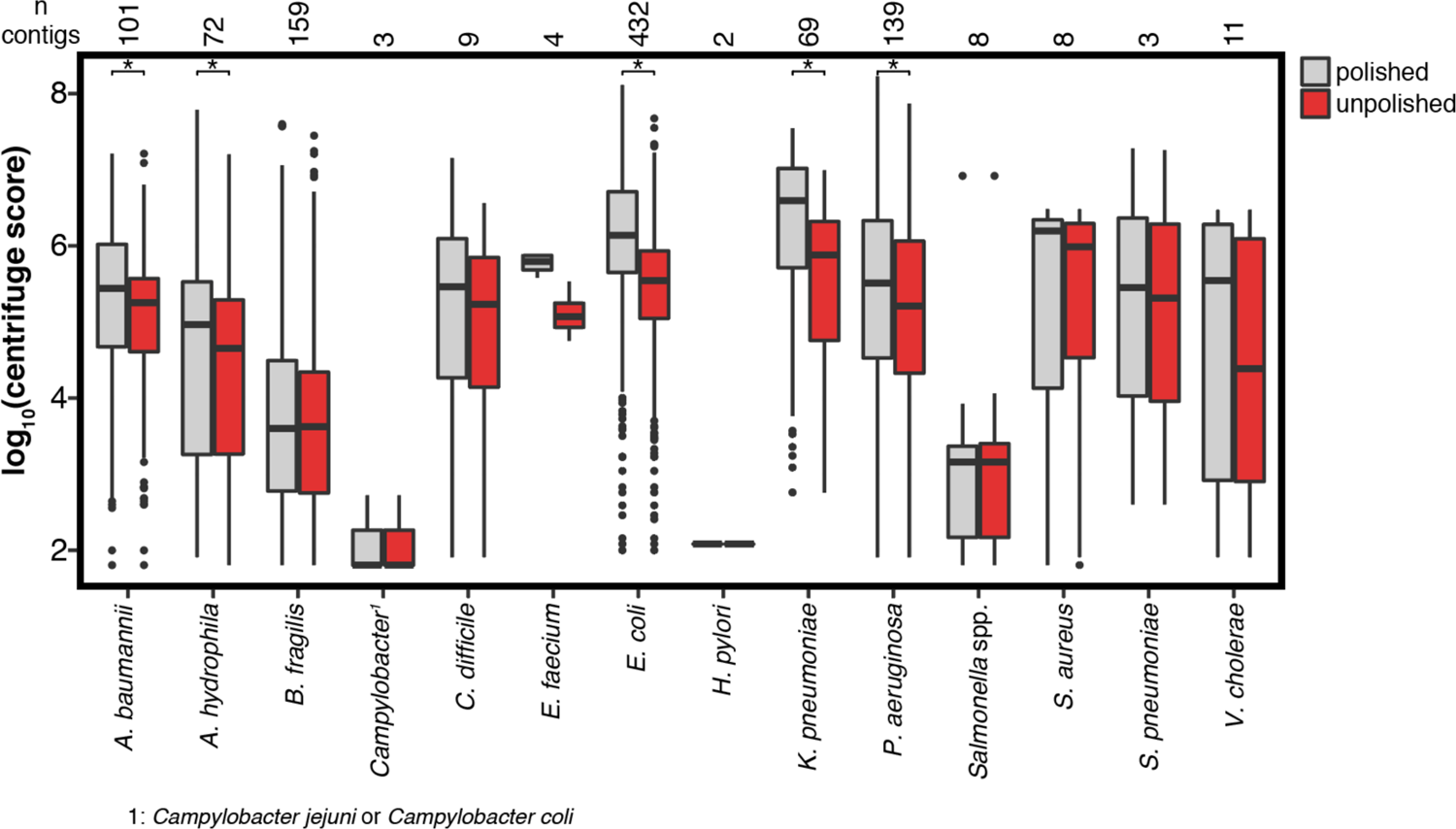
Log transformed centrifuge scores, sum of squared lengths of matched segments, of paired contigs classified as potential pathogens before and after short-read polishing. Significant differences after correcting for multiple comparisons (Bonferroni) are indicated by *. The line through each box denotes the median centrifuge score, the lower and upper box boundaries denote the first and third quartiles, respectively, and the whiskers extend to the minimum and maximum scores up to 1.5 times the interquartile range, with outliers plotted as dots.

### Identification of Antibiotic-Resistant Pathogens and Antibiotic Resistance Genes

By real-time PCR, Boston samples were positive for, on average, 53 ± 17 out of the 82 possible antibiotic resistance gene targets whereas Vellore samples were positive for 60 ± 6 antibiotic resistance genes (Table S8). Using real-time PCR as a benchmark, raw long-read metagenomic sequencing was 16% [stdev: 23%] sensitive and 100% [0%] specific at detecting clinically relevant resistance genes (Figure 1, and Table S8). It is notable that despite < 100% sequencing accuracy, long-read sequencing did not produce any false positives in the subset of 28 clinically-relevant resistance genes.

The overall abundance of resistance genes was highest in the hospital effluent samples (Vellore hospital: 4260 ARGs/Gbp; Boston hospital: 2114 ARGs/Gbp (Table S4)). 8% of ARGs in the Boston hospital effluent mapped to colistin resistance genes (Figure S3). Specifically, reads aligned to the *mcr*-*3* gene and the reads were classified as *Aeromonas* spp., which is consistent with other studies ^41, 42^. *Aeromonas* spp. carrying *mcr*-*3* genes have been found in humans, retail meat, and the environment in China and in wastewater in Japan ^41, 42^.

In the polished contigs classified as *A. baumannii* and *P. aeruginosa* we identified aminoglycoside resistance genes in Boston wastewater, surface, and hospital samples (Figure 3a). Contigs were also identified as *C. difficile* containing tetracycline resistance genes in the Boston hospital effluent and contigs classified as *S. aureus* with lincosamide resistance genes were found in Boston wastewater. In Vellore, all but one surface sample contained contigs classified as potential pathogens with resistance genes that span numerous drug classes (Figure 3a). Clinically relevant resistance genes found in pathogens included *armA* in *A. baumannii* and *bla*CTX-M in *E. coli* in the hospital wastewater in Vellore.

**Figure 3:**
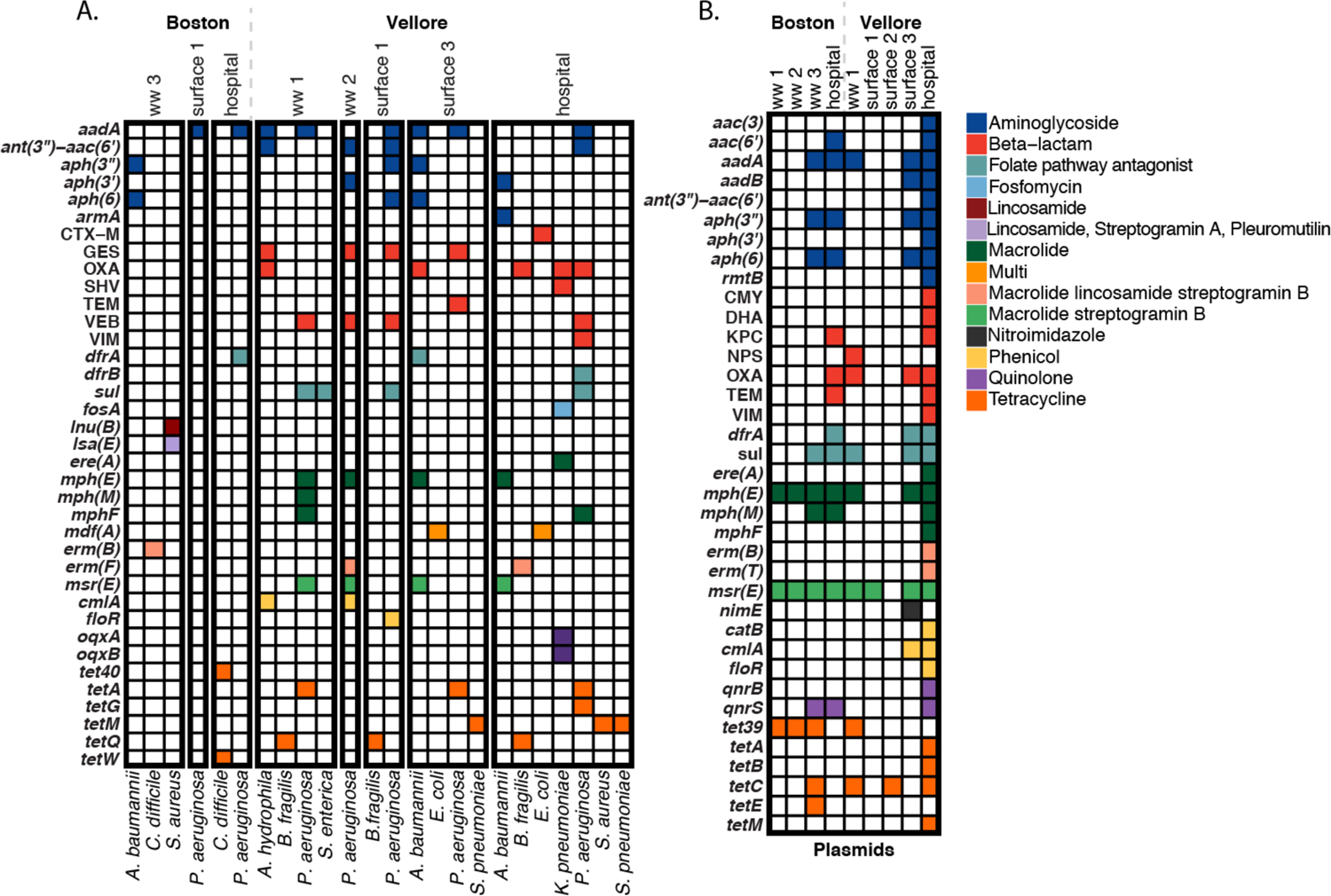
Putative antibiotic resistance genes in contigs classified as A.) potential pathogens and B.) plasmids from municipal wastewater (ww), hospital wastewater (hospital), and surface water (surface) in Boston, USA and Vellor, India. Drug class of resistance gene is indicated by color. Resistance genes were identified by aligning assembled contigs to the ResFinder database using Minimap2. Taxonomy was assigned with Centrifuge using the RefSeq database and MOB-suite was used for plasmid identification. Samples not included in the figure did not contain antibiotic resistance genes in contigs classified as potential pathogens or plasmids.

Were detected numerous resistance genes, from different classes of antibiotics, on plasmids in both Boston and Vellore (Figure 3b). Beta-lactamase genes were present on plasmids in the hospital effluents from both cities in addition to wastewater and surface samples in Vellore. It is important to note that beta-lactamase genes vary in clinical significance at the allele level, and therefore only some are extended spectrum beta-lactamases (ESBLs), conferring resistance to antibiotics of last resort. For example, the primary alignment for a *bla*_TEM_ gene found on a plasmid in the Boston hospital effluent was *bla*_TEM-6_ which is an ESBL, but the primary alignment on a plasmid in the Vellore hospital effluent was *bla*_TEM-1_, which is not (supplemental data)^43^. The concordance between ARG identification at the allele level in polished and unpolished contigs was 78% for genes conferring resistance to beta-lactamases (Table S10). Among all drug classes evaluated the average concordance between polished and unpolished contigs was 86 ± 9% at the allele level and 98 ± 2% at the gene level.

## Discussion

### Utility of Assembled Long Reads

Nanopore is a relatively new technology and accordingly there are few assembly pipelines available that have been rigorously evaluated ^33, 44–47^. Recently, there has also been emphasis on the potential utility of short reads to improve long-read assemblies through polishing ^34^). While short-read polishing has been shown to improve frame-shift errors^48, 49^ and increase accuracy of error-prone long reads^34^, here we found limited impact of short-read polishing on pathogen identification or ARG classification. Previous work has also recommended the use of hybrid assemblers of both long and short reads (e.g. OPERA-MS^50^ and HybridSPAdes^51^), but in a preliminary analysis of our data, OPERA-MS produced contigs on average shorter or similar in length to our unassembled long reads (data not shown). Previous literature evaluating assemblers on mock communities has demonstrated that metaFlye, the assembler used in this work, performed well and was able to produce highly contiguous genomes^33, 45^. It is important to consider that assembler performance varies by coverage and complexity of microbial communities. For example, in an investigation of short, long, and hybrid assemblers for analysis of wastewater metagenomes, metaFlye produced the longest contigs and most co-occurrences of antibiotic resistance genes with mobile genetic elements and pathogen gene markers but was also shown to be vulnerable to misassemblies at low coverages ^52^. Canu^44^ has also been identified as a promising assembler for long reads but requires a significant amount of computing time and memory, often exceeding that available on university high-performance computer clusters with job time and memory limitations^45, 52^.

This study contributes valuable metrics (sensitivity and specificity compared to real-time PCR) for using long-read metagenomic sequencing for identification of pathogen genes in wastewater and surface waters. Previous studies using long-read sequencing of environmental water samples have explored alternative workflows that may be useful for pathogen surveillance applications, but have not validated those methods against real-time PCR for multiple pathogens. A study utilizing long-read sequencing for water quality testing in a Montana River found that long-read sequencing was able to detect pathogenic *E. coli* genes in coliform enriched cultures but not in the uncultured samples^23^. Another study evaluating long-read sequencing of the 16S rRNA gene (amplicon sequencing) in water from informal settlements in Nepal found that amplicon sequencing correlated with qPCR results for detection of *Vibrio cholera* but recommended combined use to avoid false positives^24^. Other studies have found nanopore sequencing (metagenomic and amplicon) useful for profiling spatial and temporal variation in the core communities and function of bacteria in river samples ^53, 54^.

### Occurrence of Pathogens and Antibiotic Resistance in Boston and Vellore

Overall, the samples from Vellore had more pathogen genes detected via real-time PCR and more contigs identified as potential pathogens using metagenomics. In a previous study that used a similar TaqMan array card for detecting enteric pathogen prevalence in young children in Vellore, the most prevalent bacterial pathogens were enteroaggregative *E. coli*, enteropathogenic *E. coli*, and *Campylobacter* species ^55^. Similar pathogen profiles were found in our own analysis of surface and wastewater samples in Vellore using real-time PCR. In this work, we detected *Campylobacter* at the species level in all Vellore samples, and two samples (wastewater and surface water) were positive for *C. jejuni* or *C. coli* genes by real-time PCR (Table S5). We detected few bacterial pathogens in the Boston hospital wastewater but did detect enteroaggregative *E. coli* (*aaiC*) and *S. flexneri* (clade 1-type 3 restriction enzyme) genes. We also found all pathogenic *E. coli* genes (*aaiC, aatA, aggR, eae, bfpA, estA, LT, STh, STp, stx1, stx2, or ipaH*) in all Boston wastewater samples, but it is difficult to contextualize these results because there is limited monitoring or reporting of enteric pathogens, outside of outbreak investigations, in the United States. While there are efforts by the Centers for Disease Control and Prevention to monitor waterborne illnesses, it is known that cases of diarrheal illness are significantly underreported in the United States ^56–58^.

The *bla*_NDM-1_ gene, which confers resistance to carbapenems, was found in all Vellore samples but not in Boston samples. These results suggest *bla*_NDM-1_ resistance could be widespread in India, where the gene is thought to have originated^59^, but has yet to become prevalent in Boston. It is important to note that our sample size was limited in each country. By long-read sequencing, relative abundance of resistance genes varied by drug class and exhibited location-specific trends. Boston samples were dominated by macrolide, lincosamide, streptogramin (MLS) resistance whereas Vellore samples had a higher relative abundance of aminoglycoside and beta-lactam resistance (Figure S3). A high abundance of macrolide resistance in Boston is consistent with a previous study of resistance genes around the world, that found a high relative proportion of macrolide resistance in Europe and North America^14^. A previous study of the Indian gut microbiome showed that study participants in urban areas carried resistance genes to nearly all classes of antibiotics^60^. In our study in Vellore, an urban area, contigs that were classified as potential pathogens or plasmids also carried resistance genes to most classes of antibiotics including aminoglycosides, beta-lactams, MLS, nitroimidazole, folate pathway antagonists (including sulfonamide resistance), phenicol, quinolone, and tetracycline.

In comparing resistance genes found in this study using metagenomics to resistance genes found previously in clinical isolates, we were able to identify clinically relevant resistance genes, particularly on plasmids, in Boston and Vellore. In a recent review of antibiotic resistance in hospitals in India, the most common extended spectrum beta-lactamases were CTX-M-types^61^, which we found (*bla*_CTX-M-15_) using metagenomic sequencing in *E. coli* in the Vellore hospital effluent. In the review, common carbapenem resistance genes in *E. coli* isolates were NDM and OXA48-like genes, and common beta-lactamase genes found in *K. pneumoniae* isolates in India were *bla*_TEM_, *bla*_CTX-M_, and *bla*_OXA48-like_. In our study, few beta-lactamase genes were classified on an *E. coli* contig (*bla*_CTX-M-15_) or *K. pneumoniae* contig (*bla*_OXA_), and most were found on contigs classified as plasmids. Many of the identified plasmid types (e.g. pR23, pKP3-A, pYD786-4, pKp_Goe_414-1, pFOS18) can be found in *Enterobacteriaceae*. Of the primary alignments to OXA genes on plasmids in the Vellore hospital effluent (OXA-1, OXA-164, OXA-181, OXA-392, OXA-58), one (OXA-181) was an OXA48-like allele. *E. coli* isolates in hospitals in India also contained resistance genes to other classes of antibiotics that we identified using metagenomic sequencing on plasmids in the Vellore hospital effluent, including *tetA* and *tetB*, *dfrA17*, *sul1*, and *catB3*^61^. We also identified *armA*, conferring aminoglycoside resistance, on *A. baumanii* in Vellore hospital wastewater which is in agreement with the presence of *armA* in 96.5% of isolates of *A. baumanii* from hospitals in India^61^. Unlike Vellore clinical isolates, a study of Boston hospital isolates of *Enterobacteriaceae* found that most contained *bla*_KPC_ resistance, specifically KPC-2, KPC-3, and KPC-4^62^. We identified KPC-2 resistance genes on plasmids in both the Boston and Vellore hospital effluents, but KPC resistance is thought to be less common in India^61^.

It should be noted that our results are likely a conservative estimate of the utility of long-read sequencing, and achieving more sequencing depth through increased flow cell yields (number of base pairs sequenced) could generate even more contigs classified as pathogens or pathogens containing resistance genes. Removing shorter DNA fragments through size selection is one method that can be employed to increase flow-cell yields since shorter fragments are preferentially sequenced. In addition, identification of pathogens with sequencing is limited by the databases and methods employed to identify taxonomy. Since database limitations are difficult to overcome, we improved our ability to identify pathogens by using assembled contigs, instead of raw reads, in our pathogen analysis and used our mock community results to assess classification accuracy. Assembly increased the size of fragments (mean N_50_ reads=3462 bp, mean N_50_ contigs=47400 bp, excluding the mock community) although it is notable that the assembly for the mock community produced significantly longer contigs than the environmental samples (mock N_50_= 2.9 Mbp). Lastly, resistance genes identified using sequencing methods, without culturing and antibiotic susceptibility testing, are putative and do not guarantee phenotypic resistance.

### Wastewater Surveillance

Wastewater-based epidemiology has gained major momentum in the past year in response to COVID-19 as it provides the ability to monitor for SARS-CoV-2 RNA at the population level^10^. Our study results are timely for informing how SARS-CoV-2 wastewater monitoring could be expanded to include surveillance of other pathogens and antibiotic resistance genes. In addition, building-level surveillance has recently been implemented for identifying COVID-19 hotspots and similar sampling scales could be useful for monitoring antibiotic resistance, such as at hospitals^63^. Using long-read sequences we identified mobile resistance genes to most drug classes of antibiotics in the hospital wastewater in Vellore. Previous surveillance of hospital effluent in India has led to discovery of novel carbapenemases^26, 64^. Ongoing research during the COVID-19 pandemic to optimize sampling scales (building, sewer system, treatment plant)^65, 66^ and method of sampling (composite, grab, moore swab)^67, 68^ will also be applicable for wastewater sampling for enteric pathogens and antibiotic resistance.

## Data Availability

Sequencing data is available in the SRA under Bioproject PRJNA672704

## Acknowledgements

We thank Jie Liu for her assistance with the array card for pathogens. We would also like to acknowledge support from the Tufts Institute for the Environment. E.R.F was supported by the NSF Postdoctoral Research Fellowships in Biology Program under Grant No. 1906957. Any opinions, findings, and conclusions or recommendations expressed in this material are those of the author(s) and do not necessarily reflect the views of the National Science Foundation. L.E.V and A.M. were supported by the Tufts-CMC Framework Program for Global Health Innovation D43 TW009377 from the Fogarty International Center, National Institutes of Health. The project described was also supported by the National Center for Advancing Translational Sciences, National Institutes of Health, Award Number UL1TR002544. The content is solely the responsibility of the authors and does not necessarily represent the official views of the NIH. The authors acknowledge the Tufts University High Performance Compute Cluster (https://it.tufts.edu/high-performance-computing) which was utilized for the research reported in this paper.

## Supporting Information

### Sample Collection, Processing, and DNA Extraction

For each sample, 2.5L of water was collected with a disposable 100mL Dippas Sampling Dipper to fill five-500mL Whirl Pak bags (Nasco, Modesto, CA). Samples were placed in a cooler with ice packs, transported to the site’s laboratory facility, and filtered (Boston samples at Tufts Medical Center and Vellore samples at Christian Medical College). Filtration was conducted using a Millipore filtration unit and 0.22 µm Nanoceram disc filters (Argonide, Sanford, FL). Due to high turbidity, filters were changed when flow rates were negligible. Filters were stored at −80 °C until DNA extraction.

The DNeasy PowerWater kit (Qiagen, Germantown, MD) was used for DNA extraction according to the manufacturer’s instructions with one modification. To accommodate the large number of filters per sample, all filters for one sample were soaked in PBS solution for 30 minutes followed by perturbation of each filter surface. The supernatant was decanted and retained for bead beating. One extraction blank was included per 6 samples. Following extraction, the concentration and purity of the DNA was assessed by spectrophotometry (NanoDrop ND-200) at each laboratory. Extractions were conducted by the same lab technician across both labs. The extracted DNA was stored at −20 °C until downstream processing.

### Molecular Analyses

Each real-time PCR TAC Pathogen array card held viral, bacterial, fungal, protozoan, and helminth targets (Table S2) and could hold 8 samples. For each sample, 20 µl of sample was combined with 50 µl RT-PCR buffer (AgPathID One-Step RT-PCR Kit, Applied Biosystems), 4 µl of enzyme mix, and 6 µl of nuclease free water for a total volume of 100 µl. Each sample was mixed and deposited into the sample port on the card. The card was centrifuged twice at 1,200 rpm for 1 minute, then sealed, and the sample ports removed. The card was then placed into the thermal cycler (QuantStudio 12K Flex, Applied Biosystems) and run under the following cycling conditions: 45**°**C for 20 minutes, 95°C for 10 minutes, and 45 cycles of 95°C for 15 seconds followed by 60°C for 1 minute. One extraction blank was run as part of the 8 samples and the card contained two internal positive controls.

Each real-time PCR antibiotic resistance (AR) array card held 83 different ARG targets (Table S3) plus positive controls. One sample was used per card plus one extraction blank was also analyzed (n=13). For each sample, 500 ng of DNA was combined with 1275 µl qPCR mastermix (Microbial qPCR Mastermix, Qiagen) and a sufficient amount of nuclease free water to reach a total volume of 2550 µl. 25 µl of sample was then added to each well of the array card. Each card was run in a thermal cycler (StepOne Plus, Applied Biosystems) with the following cycling conditions: 95 °C for 10 minutes followed by 40 cycles of 95 °C for 15 seconds and 60 °C for 2 minutes. Manufacturer’s recommendations for baseline and threshold values were used; 20 cycles and 0.2, respectively.

### Mock Sample

A mock sample was developed by the CMC laboratory and included Pseudomonas aeruginosa, Methicillin-resistant Staphylococcus aureus, Klebsiella pneumoniae, Escherichia coli, Stenotrophomonas maltophilia, and Burkholderia pseudomultivorans. Each bacterial strain in the mock community had a set of known ARGs (listed in Table S1). The set of pathogenic bacterial strains used included a mix of gram positive and gram-negative bacteria, varying levels of GC content, varying genome sizes, and varying abundances of the microorganisms within the mock sample. Five of the six bacterial pathogens contained antimicrobial resistance genes.

---

**Figure S1:**
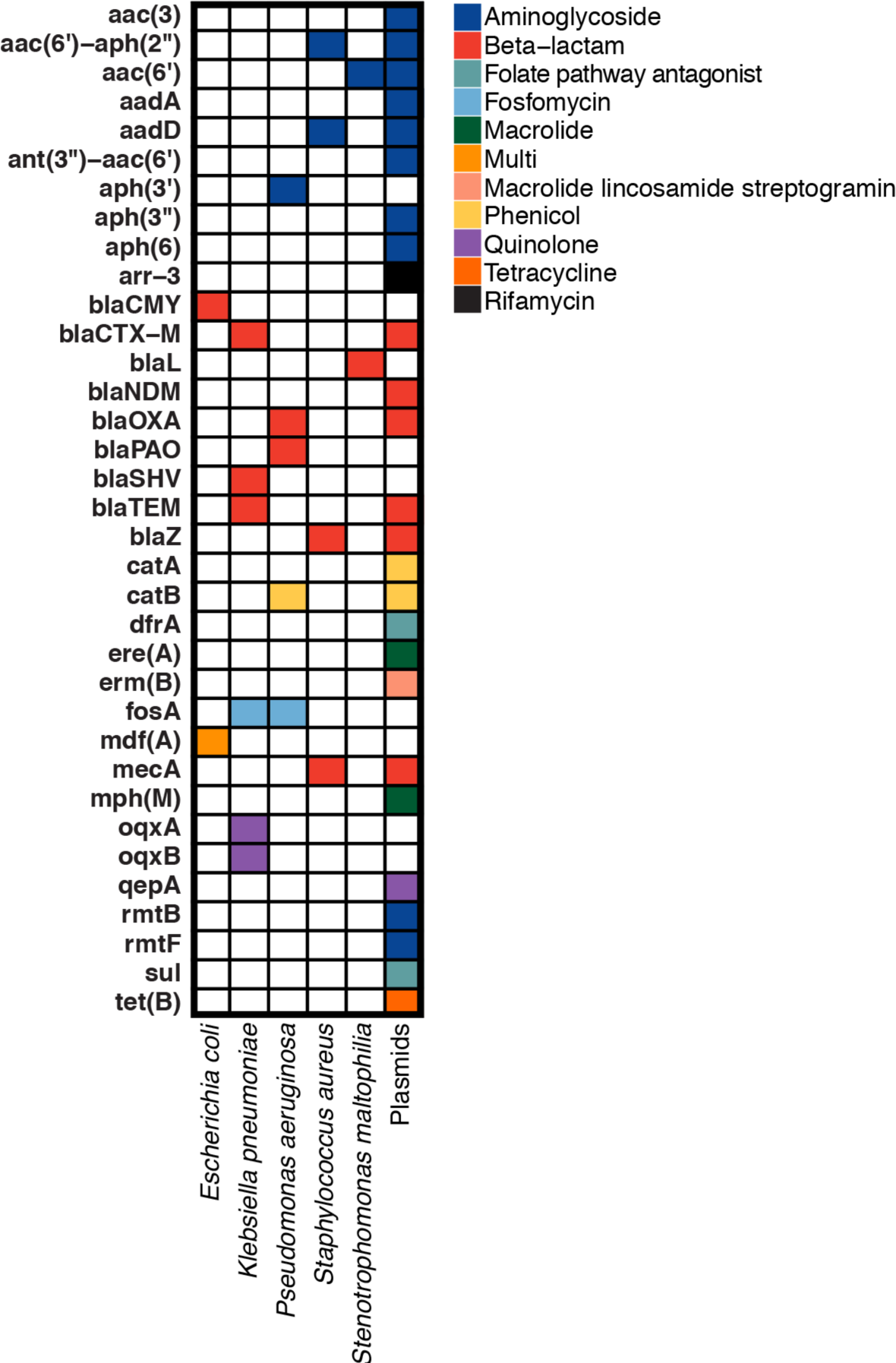
Taxonomic classification and AMR genes in contigs assembled from long-read sequences in the mock community.

**Figure S2:**
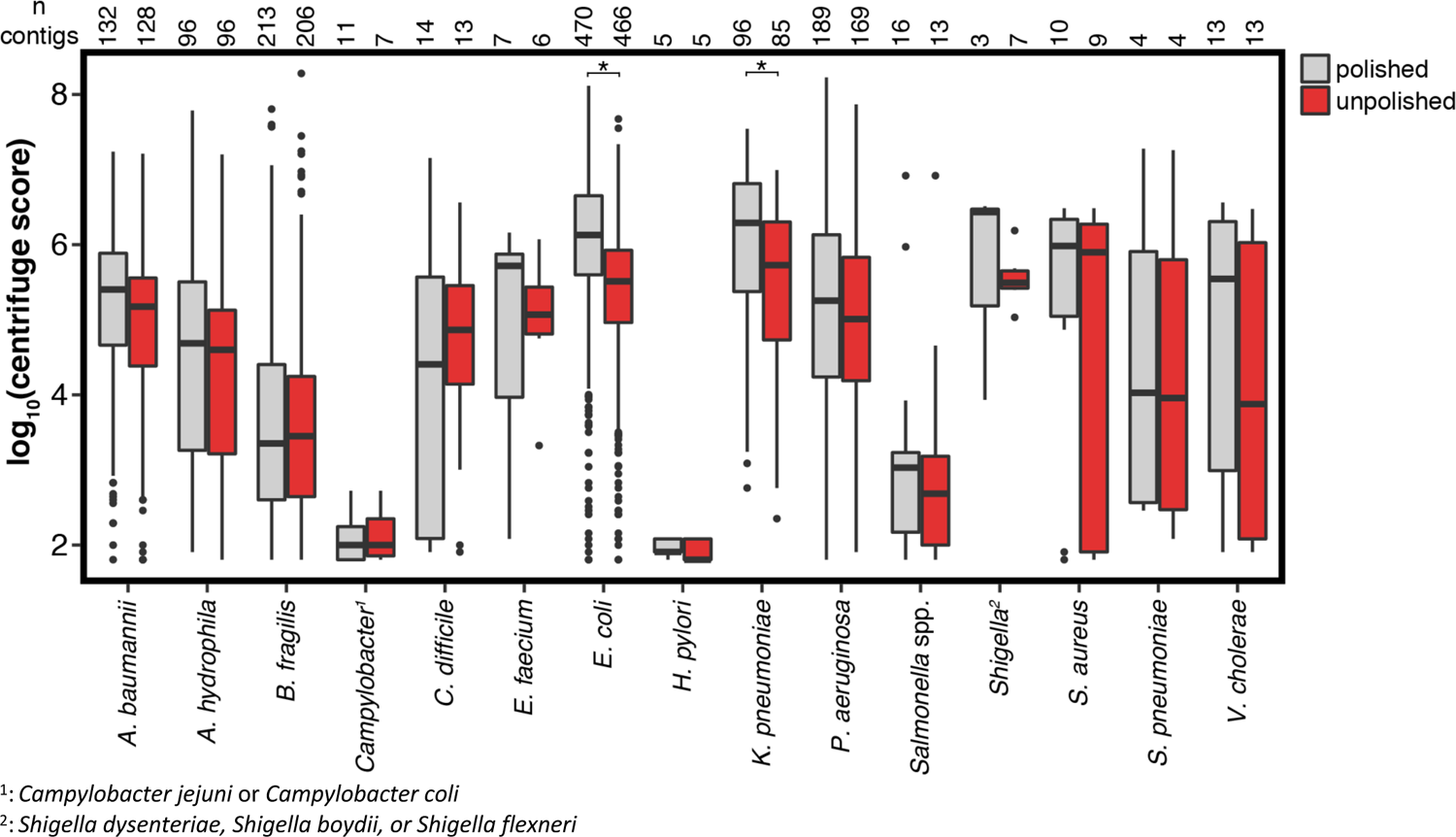
Log transformed centrifuge scores, sum of squared lengths of matched segments, of unpaired contigs classified as potential pathogens before and after short-read polishing. Significant differences after correcting for multiple comparisons are indicated by *. The line through each box denotes the median centrifuge score, the lower and upper box boundaries denote the first and third quartiles, respectively, and the whiskers extend to the minimum and maximum scores up to 1.5 times the interquartile range, with outliers plotted as dots.

**Figure S3:**
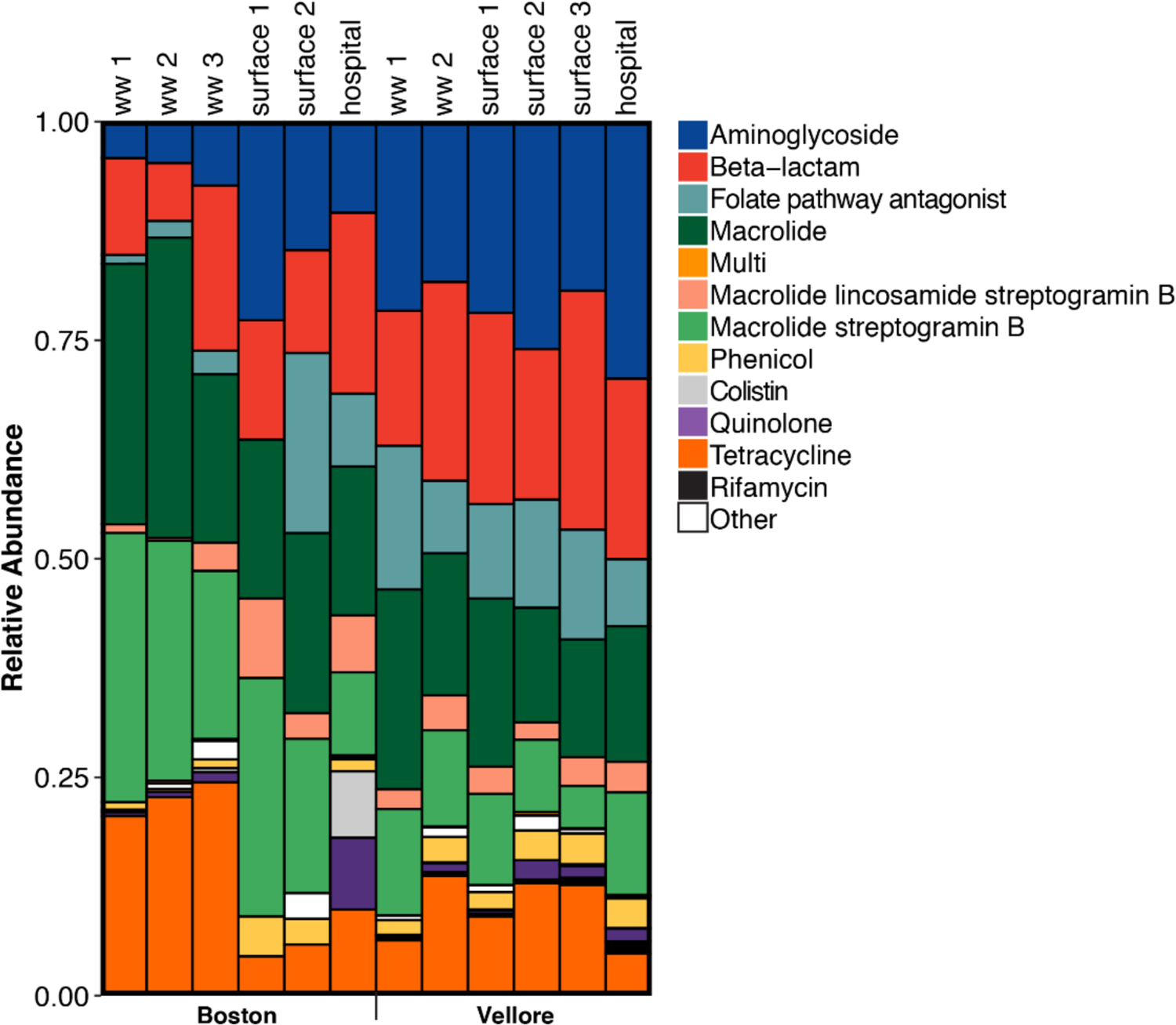
Relative abundance of resistance genes by drug class using raw long-reads.

**Table S1:** Mock community constituents and AMR genes identified in assembled contigs from long-read sequences.

| **Bacteria** | **Expected AMR Gene Targets** | **AMR Genes in Assembled Contigs** |
| --- | --- | --- |
| *Pseudomonas aeruginosa* | *OXA-10, OXA-50, VEB-1, PAO, aadA1, aph(3')-Iib, fosA, sul1, tet(A), catB7, dfrB2* | *aph(3')-Iib, blaOXA-494, blaPAO, fosA, catB7* |
| Methicillin-resistant *Staphylococcus aureus* | *blaZ, mecA, ant(6)-Ia, aph(3'')-III, acc(6')-aph(2''), ermA, norA, tet(M)* | *aac(6')-aph(2''), aadD, blaZ, mecA* |
| *Klebsiella pneumoniae* | *aadA2, aacA4, blaTEM-1B, blaSHV-1, blaCTX-M 15, blaOXA 232, oqxA, oqxB, aac(6') Ib-cr, qepA, fosA, mphA, ermB, catA1, ARR2, sul1, tetB, dfrA12* | *blaTEM-1B, blaCTX-M-15, blaSHV-106, fosA, oqxA, oqxB* |
| *Escherichia coli* | Carbapenemases, CarbaNP positive | *blaCMY-2, mdf_A* |
| *Stenotrophomonas maltophilia* | *aac(6')-Iz, aph(3')-Iic, blaL1/2, smqnr44* | *blaL1, aac(6')-Iak* |
| *Burkholderia pseudomultivorans* | None |  |
| Plasmids | None specified | *blaCTX-M-15, blaTEM-1B, blaNDM-5, blaOXA-1, blaOXA-181, ant(3'')-Ii, ARR-3, aadA2, rmtB, dfrA12, dfra17, ant(3'')-Ia, rmtF, mph_A, sul1, sul2, aph(3'')-Ib, aadA5, aac(6'')-Ib, tet(B), catA1, catB3, ant(3'')-Ia, aac(3')-IId, aph(6')-Id, qepA4, ermB, ereA* |

**Table S2:**
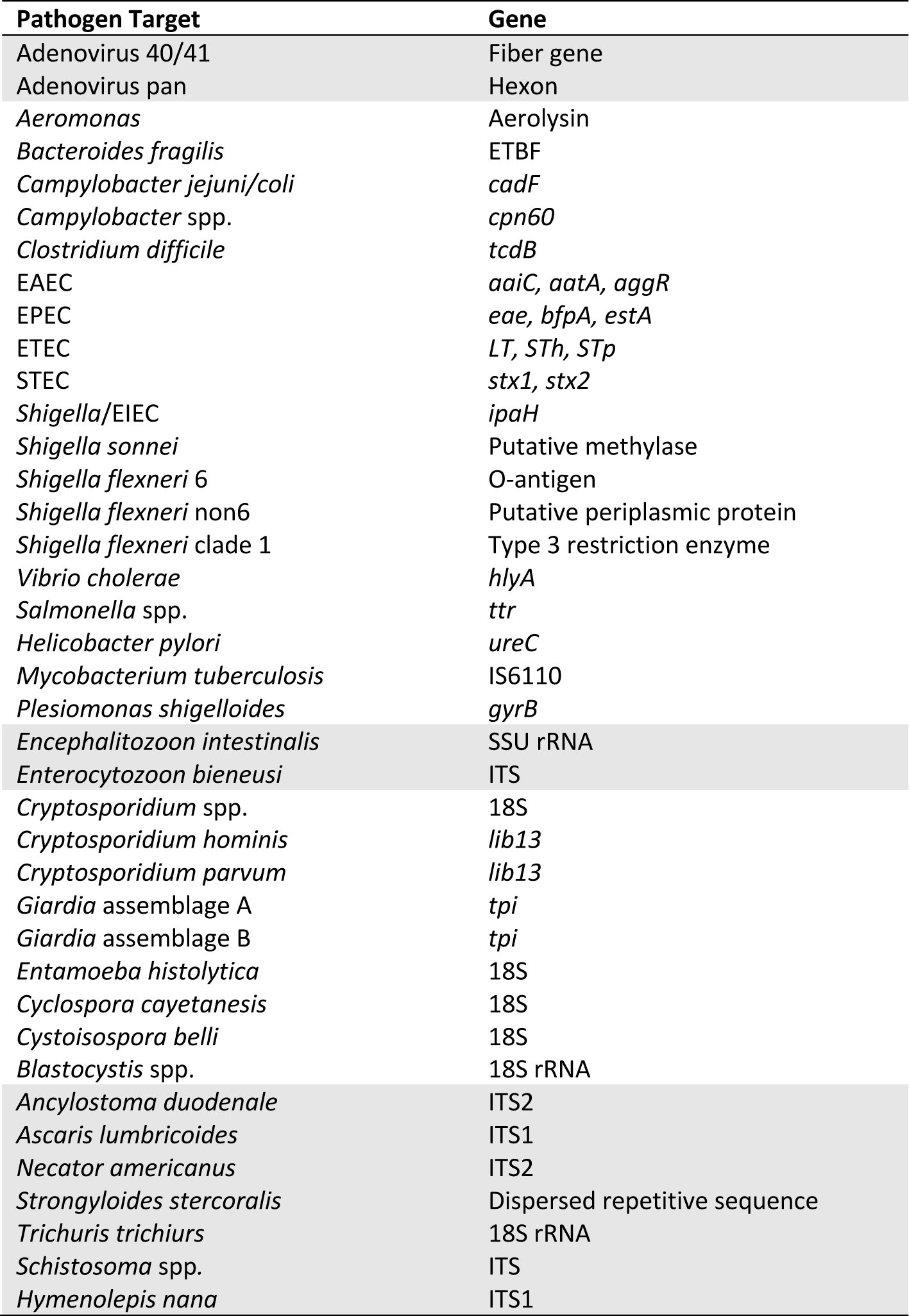
Pathogen gene targets on TAC array card.

| **Pathogen Target** | **Gene** |
| --- | --- |
| Adenovirus 40/41 | Fiber gene |
| Adenovirus pan | Hexon |
| *Aeromonas* | Aerolysin |
| *Bacteroides fragilis* | ETBF |
| *Campylobacter jejuni/coli* | *cadF* |
| *Campylobacter* spp. | *cpn60* |
| *Clostridium difficile* | *tcdB* |
| EAEC | *aaiC, aatA, aggR* |
| EPEC | *eae, bfpA, estA* |
| ETEC | *LT, ST<sub>H</sub>, ST<sub>p</sub>* |
| STEC | *stx1, stx2* |
| *Shigella*/EIEC | *ipaH* |
| *Shigella sonnei* | Putative methylase |
| *Shigella flexneri* 6 | O-antigen |
| *Shigella flexneri* non6 | Putative periplasmic protein |
| *Shigella flexneri* clade 1 | Type 3 restriction enzyme |
| *Vibrio cholerae* | *hlyA* |
| *Salmonella* spp. | *ttr* |
| *Helicobacter pylori* | *ureC* |
| *Mycobacterium tuberculosis* | IS6110 |
| *Plesiomonas shigelloides* | *gyrB* |
| *Encephalitozoon intestinalis* | SSU rRNA |
| *Enterocytozoon bieneusi* | ITS |
| *Cryptosporidium* spp. | 18S |
| *Cryptosporidium hominis* | *lib13* |
| *Cryptosporidium parvum* | *lib13* |
| *Giardia* assemblage A | *tpi* |
| *Giardia* assemblage B | *tpi* |
| *Entamoeba histolytica* | 18S |
| *Cyclospora cayetaneis* | 18S |
| *Cystoisospora belli* | 18S |
| *Blastocystis* spp. | 18S rRNA |
| *Ancylostoma duodenale* | ITS2 |
| *Ascaris lumbricoides* | ITS1 |
| *Necator americanus* | ITS2 |
| *Strongyloides stercoralis* | Dispersed repetitive sequence |
| *Trichuris trichiuris* | 18S rRNA |
| *Schistosoma* spp. | ITS |
| *Hymenolepis nana* | ITS1 |

**Table S3:** Accession number and drug class of antibiotic resistance gene targets on the real-time PCR array card.

| AR Gene Target | Accession Number | Antibiotic classification |
| --- | --- | --- |
| SFO-1 | AB003148 | Class A beta-lactamase |
| CFE-1 | AB107899 | Class C beta-lactamase |
| *qnrS* | AB187515 | Fluoroquinolone |
| *oprJ* | AE004091.2 | Multidrug resistance efflux pump |
| VEB | AF010416 | Class A beta-lactamase |
| TLA-1 | AF148067 | Class A beta-lactamase |
| SHV | AF148850 | Class A beta-lactamase |
| SHV(238S240E) | AF148851 | Class A beta-lactamase |
| GES | AF156486 | Class A beta-lactamase |
| CTX-M-9 Group | AF174129.3 | Class A beta-lactamase |
| ACC-3 | AF180958 | Class C beta-lactamase |
| CTX-M-8 Group | AF189721.1 | Class A beta-lactamase |
| *tetB* | AF223162.1 | Tetracycline efflux pump |
| BES-1 | AF234999 | Class A beta-lactamase |
| *ermB* | AF239773.1 | Macrolide Lincosamide Streptogramin_b |
| IMP-5 Group | AF290912 | Class B beta-lactamase |
| KPC | AF297554 | Class A beta-lactamase |
| CMY-10 Group | AF357597 | Class C beta-lactamase |
| OXA-24 Group | AF509241.1 | Class D beta-lactamase |
| OXA-60 | AF525303 | Class D beta-lactamase |
| OXA-23 Group | AJ132105.1 | Class D beta-lactamase |
| ACC-1 Group | AJ133121 | Class C beta-lactamase |
| IMP-2 Group | AJ243491.1 | Class B beta-lactamase |
| OXA-51 Group | AJ309734.2 | Class D beta-lactamase |
| IMP-12 Group | AJ420864.1 | Class B beta-lactamase |
| OXA-45 | AJ519683 | Class D beta-lactamase |
| IMI & NMC-A | AJ536087 | Class A beta-lactamase |
| VIM-7 | AJ536835 | Class B beta-lactamase |
| *ccrA* | AJ831823 | Class B beta-lactamase |
| SHV(238S240K) | AJ920369 | Class A beta-lactamase |
| *qnrA* | AY070235.1 | Fluoroquinolone |
| *mefA* | AY071835.1 | Macrolide Lincosamide Streptogramin_b |
| *tetA* | AY196695.1 | Tetracycline efflux pump |
| OXA-50 Group | AY306130.1 | Class D beta-lactamase |
| OXA-55 | AY343493 | Class D beta-lactamase |
| SFC-1 | AY354402 | Class A beta-lactamase |
| OXA-54 | AY500137 | Class D beta-lactamase |
| *vanB* | AY655710.1 | Vancomycin resistance |
| OXA-58 Group | AY665723.1 | Class D beta-lactamase |
| MOX | D13304 | Class C beta-lactamase |
| *msrA* | DQ068449.1 | Macrolide Lincosamide Streptogramin_b |
| *aacC4* | DQ241380 | Aminoglycoside |
| *aac(6')-Ib-cr* | DQ303918.1 | Fluoroquinolone |
| QnrB-5 Group | DQ303919.1 | Fluoroquinolone |
| QnrB-4 Group | DQ303921.2 | Fluoroquinolone |
| QnrB-1 Group | DQ351241.1 | Fluoroquinolone |
| VIM-13 | DQ365886 | Class B beta-lactamase |
| *aacC1*, *aadA1* | DQ370505.1 | Aminoglycoside |
| *aacC2* | EU022314 | Aminoglycoside |
| QnrB-8 Group | EU043312.2 | Fluoroquinolone |
| *vanC* | EU151768.1 | Vancomycin |
| *qnrC* | EU917444.1 | Fluoroquinolone |
| *aphA6* | EU977572.1 | Aminoglycoside |
| *qepA* | FJ183463.1 | Fluoroquinolone |
| *qnrD* | FJ228229.1 | Fluoroquinolone |
| ACT 5/7 Group | FJ237369 | Class C beta-lactamase |
| *ermA* | FJ390057.1 | Macrolide Lincosamide Streptogramin_b |
| SHV(156D) | FJ685656.1 | Class A beta-lactamase |
| NDM-1 | FN396876 | Class B beta-lactamase |
| BIC-1 | GQ260093 | Class A beta-lactamase |
| SHV(238G240K) | GU064396 | Class A beta-lactamase |
| OXA-48 Group | HM755942.1 | Class D beta-lactamase |
| QnrB-31 Group | HQ418999.2 | Fluoroquinolone |
| DHA | HQ456945 | Class C beta-lactamase |
| *mecA* | KC243783.1 | Beta-lactam resistance |
| *oprM* | L11616.1 | Multidrug resistance efflux pump |
| MIR | M37839.2 | Class C beta-lactamase |
| IMP-1 Group | S71932.1 | Class B beta-lactamase |
| ACT-1 Group | U58495 | Class C beta-lactamase |
| OXA-18 | U85514 | Class D beta-lactamase |
| *lukF* | VFG1276 | Panton-Valentine leukocidin chain F precursor |
| *ereB* | X03988.1 | Erythromycin resistance |
| OXA-2 Group | X07260.1 | Class D beta-lactamase |
| FOX | X77455 | Class C beta-lactamase |
| LAT | X78117 | Class C beta-lactamase |
| CTX-M-1 Group | X92506 | Class A beta-lactamase |
| Per-2 Group | X93314 | Class A beta-lactamase |
| *ermC* | Y09002.1 | Macrolide Lincosamide Streptogramin_b |
| VIM-1 Group | Y18050.2 | Class B beta-lactamase |
| Per-1 Group | Z21957.1 | Class A beta-lactamase |
| OXA-10 Group | Z22590 | Class D beta-lactamase |
| SME | Z28968 | Class A beta-lactamase |
| *S. aureus* | NZ_ACOT01000039.1 | *Staphylococcus aureus*, *Staphylococcus epidermidis* |
| *spa* | VFG1313 | *Staphylococcus aureus* |

**Table S4:** Illumina and Nanopore sequencing summary statistics. F=forward reads, R=reverse reads, Ww=wastewater.

| Illumina |  |  |  | Nanopore |  |  |  |  |
| --- | --- | --- | --- | --- | --- | --- | --- | --- |
|  | Sample | No. Illumina Reads<br>(Paired end-in millions) | Gbp Illumina Sequenced<br>(F and R) | Gbp Nanopore Sequenced | N50 Nanopore Reads | N50 Contigs Kbp<br>(unpolished) | N50 Contigs Kbp<br>(polished) | Total Abundance of Resistance Genes (per Gbp of bacterial species) |
| Boston | Ww 1 | 19.30 | 5.68 | 1.32 | 5143 | 33.1 | 33.1 | 606 |
|  | Ww 2 | 18.44 | 5.45 | 0.82 | 5749 | 28.2 | 28.2 | 444 |
|  | Ww 3 | 16.79 | 4.92 | 5.03 | 6044 | 38.3 | 38.2 | 555 |
|  | Surface 1 | 20.16 | 5.92 | 7.21 | 1997 | 21.0 | 20.9 | 6 |
|  | Surface 2 | 18.89 | 5.39 | 2.26 | 1341 | 36.5 | 36.6 | 31 |
|  | Hospital | 26.68 | 7.78 | 2.21 | 2151 | 60.6 | 60.3 | 2114 |
| Vellore | Ww 1 | 22.85 | 6.57 | 3.81 | 4547 | 70.8 | 70.7 | 953 |
|  | Ww 2 | 25.25 | 7.18 | 4.23 | 2152 | 58.8 | 58.7 | 1500 |
|  | Surface 1 | 26.77 | 7.76 | 2.55 | 2313 | 58.0 | 57.8 | 1454 |
|  | Surface 2 | 32.50 | 9.14 | 2.13 | 986 | 40.3 | 40.1 | 695 |
|  | Surface 3 | 21.56 | 6.12 | 3.94 | 4284 | 64.8 | 64.8 | 1192 |
|  | Hospital | 24.31 | 6.96 | 5.67 | 4836 | 58.3 | 58.1 | 4260 |
|  | Mock | NA | NA | 0.96 | 8199 | 2885 | NA | NA |

**Table S5:** Presence (1) and absence (0) of pathogen gene targets in Boston and Vellore by TAC. Ww=wastewater.

| Target | Boston |  |  |  |  |  | Vellore |  |  |  |  |  |
| --- | --- | --- | --- | --- | --- | --- | --- | --- | --- | --- | --- | --- |
|  | Ww 1 | Ww 2 | Ww 3 | Surface 1 | Surface 2 | Hospital | Ww 1 | Ww 2 | Surface 1 | Surface 2 | Surface 3 | Hospital |
| Adenovirus 40/41 | 1 | 1 | 1 | 1 | 1 | 0 | 1 | 1 | 1 | 1 | 1 | 1 |
| Adenovirus pan | 1 | 1 | 1 | 1 | 1 | 0 | 1 | 1 | 1 | 1 | 1 | 1 |
| *Aeromonas* | 1 | 1 | 1 | 1 | 1 | 1 | 1 | 1 | 1 | 1 | 1 | 1 |
| *Bacteroides fragilis* | 1 | 1 | 1 | 0 | 1 | 1 | 1 | 1 | 1 | 1 | 1 | 1 |
| *Campylobacter jejuni/coli* | 0 | 0 | 0 | 0 | 0 | 0 | 0 | 1 | 1 | 0 | 0 | 0 |
| *Campylobacter* spp. | 1 | 1 | 1 | 1 | 1 | 1 | 1 | 1 | 1 | 1 | 1 | 1 |
| *Clostridium difficile* | 0 | 1 | 1 | 0 | 0 | 0 | 0 | 0 | 0 | 0 | 0 | 1 |
| EAEC aaiC | 1 | 1 | 1 | 0 | 1 | 1 | 1 | 1 | 1 | 1 | 1 | 1 |
| EAEC aatA | 1 | 1 | 1 | 1 | 1 | 0 | 1 | 1 | 1 | 1 | 1 | 1 |
| EAEC aggR | 1 | 1 | 1 | 1 | 1 | 0 | 1 | 1 | 1 | 1 | 1 | 1 |
| EPEC bfpA | 1 | 1 | 1 | 0 | 0 | 0 | 1 | 1 | 1 | 0 | 1 | 1 |
| EPEC eae | 1 | 1 | 1 | 0 | 1 | 0 | 1 | 1 | 1 | 1 | 1 | 1 |
| ETEC LT | 1 | 1 | 1 | 0 | 0 | 0 | 1 | 1 | 1 | 1 | 1 | 1 |
| ETEC STp | 1 | 1 | 1 | 0 | 0 | 0 | 1 | 1 | 1 | 1 | 1 | 1 |
| STEC stx1 | 1 | 0 | 1 | 0 | 0 | 0 | 0 | 1 | 0 | 1 | 1 | 1 |
| STEC stx2 | 0 | 1 | 1 | 0 | 0 | 0 | 1 | 1 | 1 | 0 | 1 | 1 |
| *Shigella*/EIEC | 1 | 1 | 1 | 0 | 0 | 0 | 1 | 1 | 1 | 1 | 1 | 1 |
| *Shigella sonnei* | 0 | 1 | 0 | 0 | 0 | 0 | 0 | 0 | 0 | 1 | 1 | 1 |
| *Shigella flexneri* 6 | 1 | 1 | 1 | 0 | 0 | 0 | 1 | 1 | 1 | 1 | 1 | 1 |
| *Shigella flexneri* non6 | 0 | 0 | 1 | 0 | 0 | 0 | 1 | 1 | 1 | 1 | 1 | 1 |
| *Shigella flexneri* clade 1 | 1 | 1 | 1 | 1 | 1 | 1 | 1 | 1 | 1 | 1 | 1 | 1 |
| *Vibrio cholerae* | 0 | 1 | 0 | 0 | 0 | 0 | 1 | 1 | 1 | 1 | 1 | 1 |
| *Salmonella* spp. | 0 | 0 | 1 | 0 | 1 | 0 | 0 | 0 | 1 | 1 | 0 | 1 |
| *Helicobacter pylori* | 0 | 0 | 0 | 0 | 0 | 0 | 0 | 0 | 0 | 0 | 0 | 0 |
| *Mycobacterium tuberculosis* | 0 | 0 | 0 | 0 | 0 | 0 | 0 | 0 | 0 | 0 | 0 | 0 |
| *Plesiomonas shigelloides* | 0 | 1 | 1 | 1 | 1 | 0 | 1 | 1 | 1 | 1 | 1 | 1 |
| *Encephalitozoon intestinalis* | 1 | 1 | 1 | 1 | 1 | 0 | 0 | 0 | 0 | 0 | 0 | 0 |
| *Enterocytozoon bienersi* | 0 | 0 | 0 | 0 | 0 | 0 | 0 | 1 | 0 | 1 | 0 | 0 |
| *Cryptosporidium* spp. | 0 | 1 | 0 | 1 | 1 | 0 | 0 | 1 | 1 | 1 | 1 | 1 |
| *Cryptosporidium hominis* | 0 | 0 | 0 | 0 | 0 | 0 | 0 | 0 | 0 | 0 | 0 | 0 |
| *Cryptosporidium parvum* | 0 | 0 | 0 | 0 | 0 | 0 | 0 | 1 | 0 | 0 | 0 | 0 |
| *Giardia* assemblage A | 1 | 0 | 0 | 0 | 0 | 1 | 1 | 1 | 0 | 0 | 1 | 1 |
| *Giardia* assemblage B | 0 | 0 | 0 | 0 | 0 | 0 | 0 | 1 | 0 | 1 | 1 | 1 |
| *Entamoeba histolytica* | 0 | 0 | 0 | 0 | 0 | 0 | 1 | 1 | 0 | 0 | 1 | 1 |
| *Cyclospora cayetanensis* | 0 | 0 | 0 | 0 | 0 | 0 | 0 | 0 | 0 | 0 | 0 | 0 |
| *Cystoisospora belli* | 0 | 0 | 0 | 0 | 0 | 0 | 0 | 0 | 0 | 0 | 0 | 0 |
| *Blastocystis* spp. | 1 | 1 | 1 | 1 | 1 | 1 | 1 | 1 | 1 | 1 | 1 | 1 |
| *Ancylostoma duodenale* | 0 | 0 | 0 | 0 | 0 | 0 | 0 | 0 | 0 | 0 | 0 | 0 |
| *Ascaris lumbricoides* | 0 | 0 | 0 | 0 | 0 | 0 | 0 | 0 | 0 | 0 | 0 | 0 |
| *Necator americanus* | 0 | 0 | 0 | 0 | 0 | 0 | 0 | 0 | 0 | 0 | 0 | 1 |
| *Strongyloides stercoralis* | 0 | 0 | 0 | 0 | 0 | 0 | 1 | 0 | 0 | 0 | 1 | 1 |
| *Trichuris trichiuris* | 0 | 0 | 0 | 0 | 0 | 0 | 0 | 0 | 0 | 0 | 0 | 0 |
| *Schistosoma* spp. | 0 | 0 | 0 | 0 | 0 | 0 | 0 | 0 | 0 | 0 | 0 | 0 |
| *Hymenolepis nana* | 0 | 0 | 0 | 0 | 0 | 0 | 0 | 0 | 0 | 0 | 0 | 0 |

**Table S6:** Conditional probabilities comparing polished and unpolished contigs to real-time PCR for pathogen identification.

| Pathogen | Polished |  |  |  | Unpolished |  |  |  |
| --- | --- | --- | --- | --- | --- | --- | --- | --- |
|  | Sensitivity | Specificity | Positive Predictive Value | Negative Predictive Value | Sensitivity | Specificity | Positive Predictive Value | Negative Predictive Value |
| *Aeromonas* | 75.00 | NA | 100.00 | 0.00 | 66.67 | NA | 100.00 | 0.00 |
| *B. fragilis* | 63.64 | 100.00 | 100.00 | 20.00 | 63.64 | 100.00 | 100.00 | 20.00 |
| *Campylobacter jejuni/coli* | 100.00 | 50.00 | 28.57 | 100.00 | 100.00 | 70.00 | 40.00 | 100.00 |
| *C. difficile* | 66.67 | 55.56 | 33.33 | 83.33 | 66.67 | 77.78 | 50.00 | 87.50 |
| *V. cholerae* | 57.14 | 100.00 | 100.00 | 62.50 | 71.43 | 80.00 | 83.33 | 66.67 |
| *Salmonella* spp. | 80.00 | 57.14 | 57.14 | 80.00 | 20.00 | 71.43 | 33.33 | 55.56 |
| *H. pylori* | NA | 66.67 | 0.00 | 100.00 | NA | 66.67 | 0.00 | 100.00 |
| EAEC | 8.33 | NA | 100.00 | 0.00 | 16.67 | NA | 100.00 | 0.00 |
| EPEC | 0.00 | 100.00 | NA | 16.67 | 40.00 | 100.00 | 100.00 | 25.00 |
| ETEC | 44.44 | 100.00 | 100.00 | 37.50 | 44.44 | 100.00 | 100.00 | 37.50 |
| Shigella/EIEC | 25.00 | NA | 100.00 | 0.00 | 33.33 | NA | 100.00 | 0.00 |
| STEC | 22.22 | 66.67 | 66.67 | 22.22 | 33.33 | 66.67 | 75.00 | 25.00 |
| *S. aureus* | 40.00 | 57.14 | 40.00 | 57.14 | 40.00 | 57.14 | 40.00 | 57.14 |

**Table S7:**
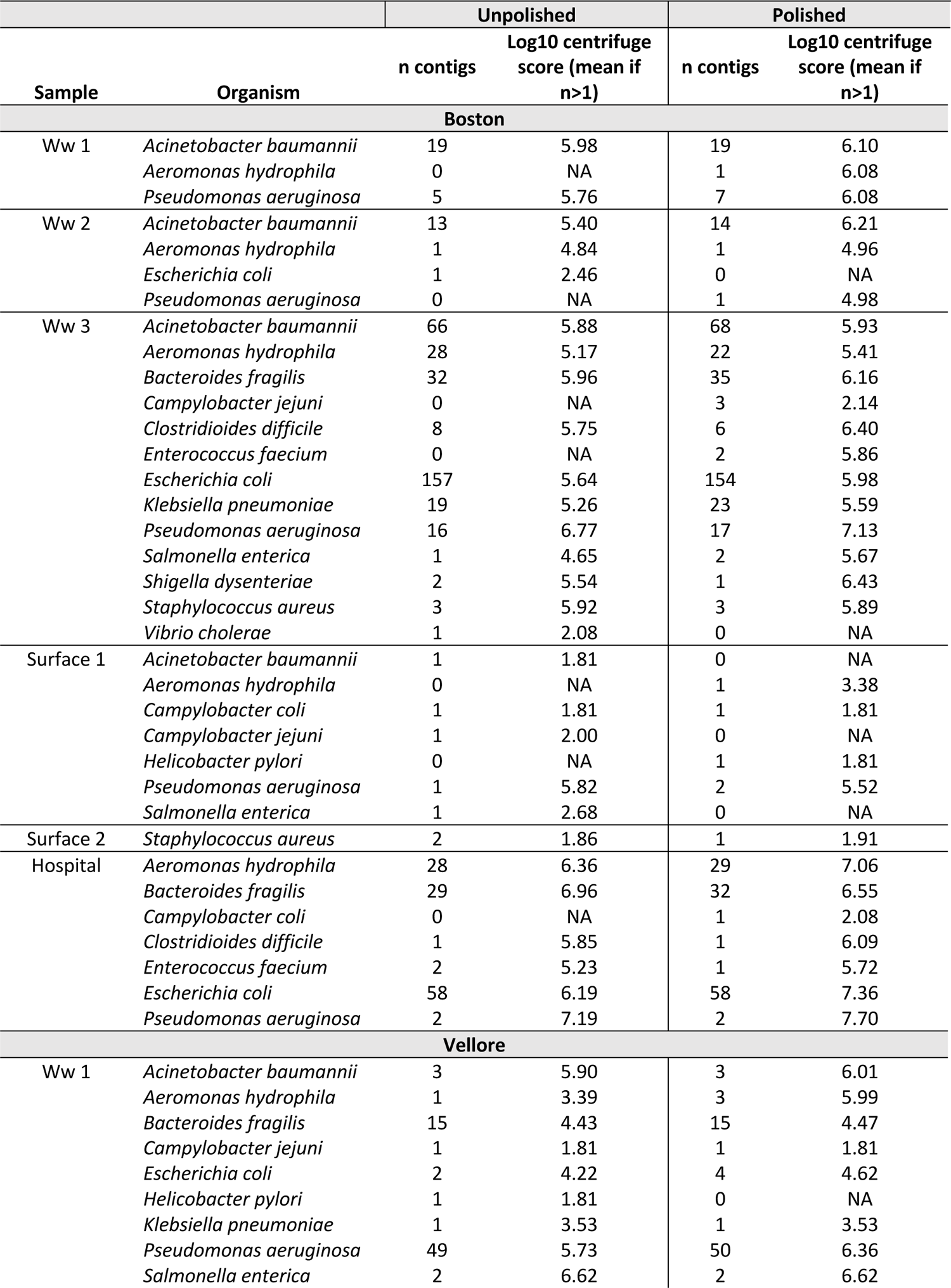
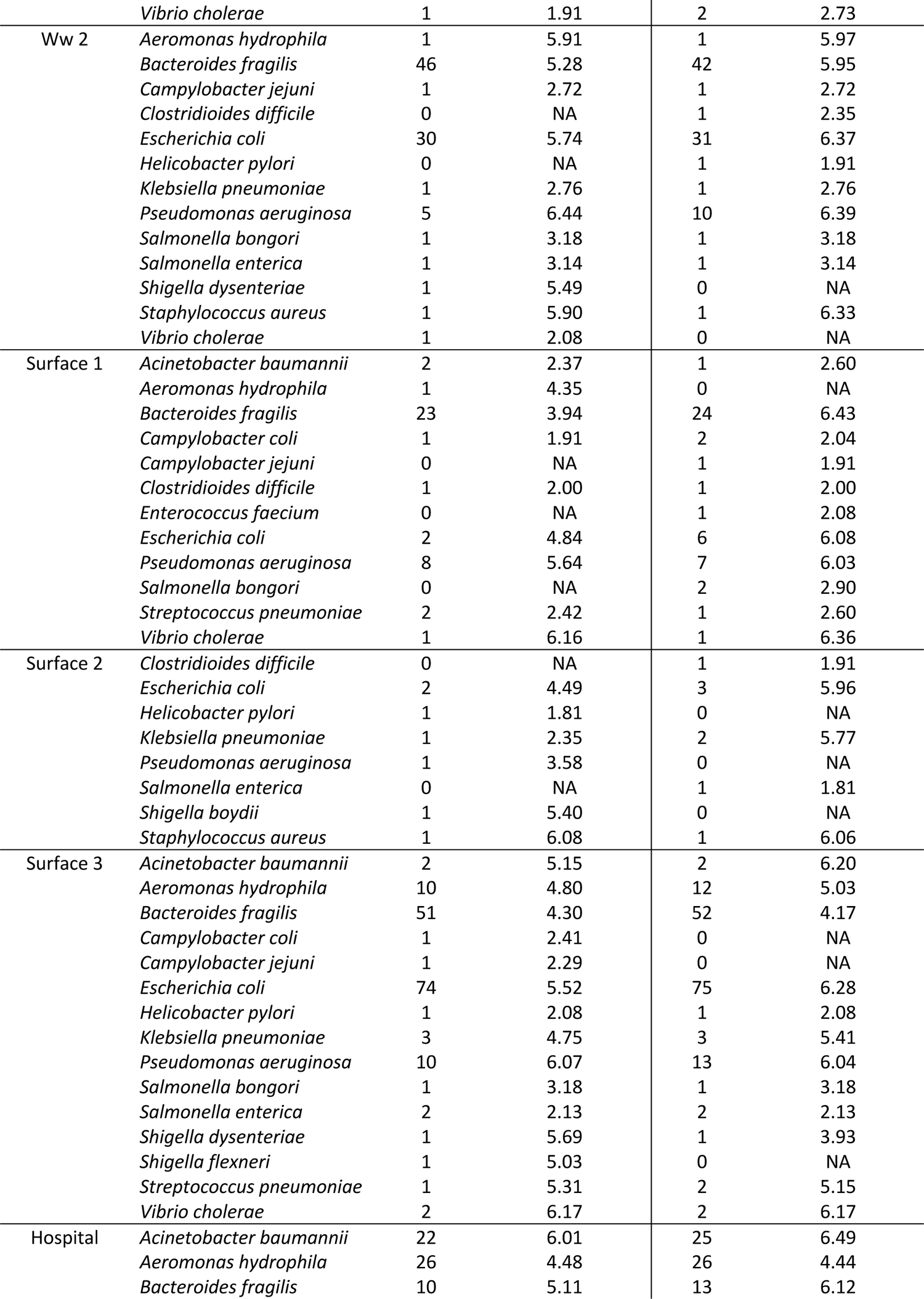
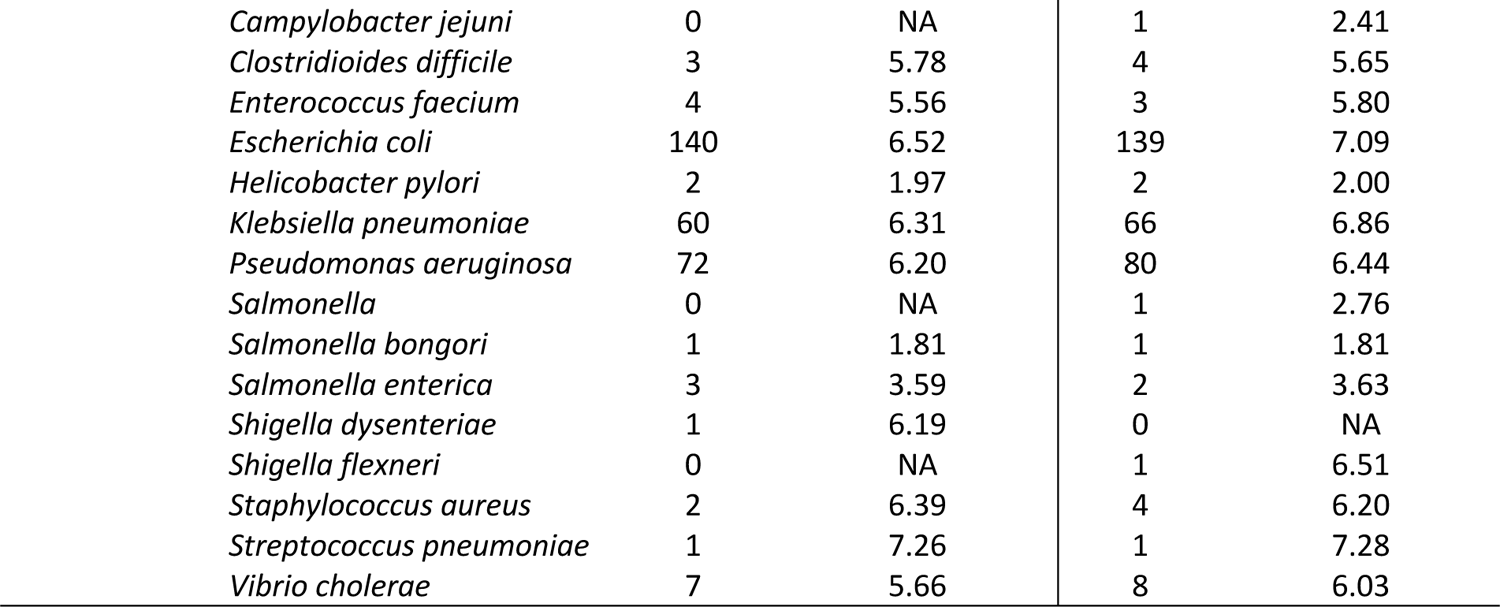
Number of contigs classified to potential pathogens and mean centrifuge score for unpolished (no short-read polishing) and polished (short-read polishing) contigs. Ww= wastewater.

|  |  | Unpolished |  | Polished |  |
| --- | --- | --- | --- | --- | --- |
| Sample | Organism | n contigs | Log10 centrifuge score (mean if n>1) | n contigs | Log10 centrifuge score (mean if n>1) |
| **Boston** |  |  |  |  |  |
| Ww 1 | *Acinetobacter baumannii* | 19 | 5.98 | 19 | 6.10 |
|  | *Aeromonas hydrophila* | 0 | NA | 1 | 6.08 |
|  | *Pseudomonas aeruginosa* | 5 | 5.76 | 7 | 6.08 |
| Ww 2 | *Acinetobacter baumannii* | 13 | 5.40 | 14 | 6.21 |
|  | *Aeromonas hydrophila* | 1 | 4.84 | 1 | 4.96 |
|  | *Escherichia coli* | 1 | 2.46 | 0 | NA |
|  | *Pseudomonas aeruginosa* | 0 | NA | 1 | 4.98 |
| Ww 3 | *Acinetobacter baumannii* | 66 | 5.88 | 68 | 5.93 |
|  | *Aeromonas hydrophila* | 28 | 5.17 | 22 | 5.41 |
|  | *Bacteroides fragilis* | 32 | 5.96 | 35 | 6.16 |
|  | *Campylobacter jejuni* | 0 | NA | 3 | 2.14 |
|  | *Clostridioides difficile* | 8 | 5.75 | 6 | 6.40 |
|  | *Enterococcus faecium* | 0 | NA | 2 | 5.86 |
|  | *Escherichia coli* | 157 | 5.64 | 154 | 5.98 |
|  | *Klebsiella pneumoniae* | 19 | 5.26 | 23 | 5.59 |
|  | *Pseudomonas aeruginosa* | 16 | 6.77 | 17 | 7.13 |
|  | *Salmonella enterica* | 1 | 4.65 | 2 | 5.67 |
|  | *Shigella dysenteriae* | 2 | 5.54 | 1 | 6.43 |
|  | *Staphylococcus aureus* | 3 | 5.92 | 3 | 5.89 |
|  | *Vibrio cholerae* | 1 | 2.08 | 0 | NA |
| Surface 1 | *Acinetobacter baumannii* | 1 | 1.81 | 0 | NA |
|  | *Aeromonas hydrophila* | 0 | NA | 1 | 3.38 |
|  | *Campylobacter coli* | 1 | 1.81 | 1 | 1.81 |
|  | *Campylobacter jejuni* | 1 | 2.00 | 0 | NA |
|  | *Helicobacter pylori* | 0 | NA | 1 | 1.81 |
|  | *Pseudomonas aeruginosa* | 1 | 5.82 | 2 | 5.52 |
|  | *Salmonella enterica* | 1 | 2.68 | 0 | NA |
| Surface 2 | *Staphylococcus aureus* | 2 | 1.86 | 1 | 1.91 |
| Hospital | *Aeromonas hydrophila* | 28 | 6.36 | 29 | 7.06 |
|  | *Bacteroides fragilis* | 29 | 6.96 | 32 | 6.55 |
|  | *Campylobacter coli* | 0 | NA | 1 | 2.08 |
|  | *Clostridioides difficile* | 1 | 5.85 | 1 | 6.09 |
|  | *Enterococcus faecium* | 2 | 5.23 | 1 | 5.72 |
|  | *Escherichia coli* | 58 | 6.19 | 58 | 7.36 |
|  | *Pseudomonas aeruginosa* | 2 | 7.19 | 2 | 7.70 |
| **Vellore** |  |  |  |  |  |
| Ww 1 | *Acinetobacter baumannii* | 3 | 5.90 | 3 | 6.01 |
|  | *Aeromonas hydrophila* | 1 | 3.39 | 3 | 5.99 |
|  | *Bacteroides fragilis* | 15 | 4.43 | 15 | 4.47 |
|  | *Campylobacter jejuni* | 1 | 1.81 | 1 | 1.81 |
|  | *Escherichia coli* | 2 | 4.22 | 4 | 4.62 |
|  | *Helicobacter pylori* | 1 | 1.81 | 0 | NA |
|  | *Klebsiella pneumoniae* | 1 | 3.53 | 1 | 3.53 |
|  | *Pseudomonas aeruginosa* | 49 | 5.73 | 50 | 6.36 |
|  | *Salmonella enterica* | 2 | 6.62 | 2 | 6.62 |
|  | *Vibrio cholerae* | 1 | 1.91 | 2 | 2.73 |
| Ww 2 | *Aeromonas hydrophila* | 1 | 5.91 | 1 | 5.97 |
|  | *Bacteroides fragilis* | 46 | 5.28 | 42 | 5.95 |
|  | *Campylobacter jejuni* | 1 | 2.72 | 1 | 2.72 |
|  | *Clostridioides difficile* | 0 | NA | 1 | 2.35 |
|  | *Escherichia coli* | 30 | 5.74 | 31 | 6.37 |
|  | *Helicobacter pylori* | 0 | NA | 1 | 1.91 |
|  | *Klebsiella pneumoniae* | 1 | 2.76 | 1 | 2.76 |
|  | *Pseudomonas aeruginosa* | 5 | 6.44 | 10 | 6.39 |
|  | *Salmonella bongori* | 1 | 3.18 | 1 | 3.18 |
|  | *Salmonella enterica* | 1 | 3.14 | 1 | 3.14 |
|  | *Shigella dysenteriae* | 1 | 5.49 | 0 | NA |
|  | *Staphylococcus aureus* | 1 | 5.90 | 1 | 6.33 |
|  | *Vibrio cholerae* | 1 | 2.08 | 0 | NA |
| Surface 1 | *Acinetobacter baumannii* | 2 | 2.37 | 1 | 2.60 |
|  | *Aeromonas hydrophila* | 1 | 4.35 | 0 | NA |
|  | *Bacteroides fragilis* | 23 | 3.94 | 24 | 6.43 |
|  | *Campylobacter coli* | 1 | 1.91 | 2 | 2.04 |
|  | *Campylobacter jejuni* | 0 | NA | 1 | 1.91 |
|  | *Clostridioides difficile* | 1 | 2.00 | 1 | 2.00 |
|  | *Enterococcus faecium* | 0 | NA | 1 | 2.08 |
|  | *Escherichia coli* | 2 | 4.84 | 6 | 6.08 |
|  | *Pseudomonas aeruginosa* | 8 | 5.64 | 7 | 6.03 |
|  | *Salmonella bongori* | 0 | NA | 2 | 2.90 |
|  | *Streptococcus pneumoniae* | 2 | 2.42 | 1 | 2.60 |
|  | *Vibrio cholerae* | 1 | 6.16 | 1 | 6.36 |
| Surface 2 | *Clostridioides difficile* | 0 | NA | 1 | 1.91 |
|  | *Escherichia coli* | 2 | 4.49 | 3 | 5.96 |
|  | *Helicobacter pylori* | 1 | 1.81 | 0 | NA |
|  | *Klebsiella pneumoniae* | 1 | 2.35 | 2 | 5.77 |
|  | *Pseudomonas aeruginosa* | 1 | 3.58 | 0 | NA |
|  | *Salmonella enterica* | 0 | NA | 1 | 1.81 |
|  | *Shigella boydii* | 1 | 5.40 | 0 | NA |
|  | *Staphylococcus aureus* | 1 | 6.08 | 1 | 6.06 |
| Surface 3 | *Acinetobacter baumannii* | 2 | 5.15 | 2 | 6.20 |
|  | *Aeromonas hydrophila* | 10 | 4.80 | 12 | 5.03 |
|  | *Bacteroides fragilis* | 51 | 4.30 | 52 | 4.17 |
|  | *Campylobacter coli* | 1 | 2.41 | 0 | NA |
|  | *Campylobacter jejuni* | 1 | 2.29 | 0 | NA |
|  | *Escherichia coli* | 74 | 5.52 | 75 | 6.28 |
|  | *Helicobacter pylori* | 1 | 2.08 | 1 | 2.08 |
|  | *Klebsiella pneumoniae* | 3 | 4.75 | 3 | 5.41 |
|  | *Pseudomonas aeruginosa* | 10 | 6.07 | 13 | 6.04 |
|  | *Salmonella bongori* | 1 | 3.18 | 1 | 3.18 |
|  | *Salmonella enterica* | 2 | 2.13 | 2 | 2.13 |
|  | *Shigella dysenteriae* | 1 | 5.69 | 1 | 3.93 |
|  | *Shigella flexneri* | 1 | 5.03 | 0 | NA |
|  | *Streptococcus pneumoniae* | 1 | 5.31 | 2 | 5.15 |
|  | *Vibrio cholerae* | 2 | 6.17 | 2 | 6.17 |
| Hospital | *Acinetobacter baumannii* | 22 | 6.01 | 25 | 6.49 |
|  | *Aeromonas hydrophila* | 26 | 4.48 | 26 | 4.44 |
|  | *Bacteroides fragilis* | 10 | 5.11 | 13 | 6.12 |

|  |  |  |  |  |
| --- | --- | --- | --- | --- |
| *Campylobacter jejuni* | 0 | NA | 1 | 2.41 |
| *Clostridioides difficile* | 3 | 5.78 | 4 | 5.65 |
| *Enterococcus faecium* | 4 | 5.56 | 3 | 5.80 |
| *Escherichia coli* | 140 | 6.52 | 139 | 7.09 |
| *Helicobacter pylori* | 2 | 1.97 | 2 | 2.00 |
| *Klebsiella pneumoniae* | 60 | 6.31 | 66 | 6.86 |
| *Pseudomonas aeruginosa* | 72 | 6.20 | 80 | 6.44 |
| *Salmonella* | 0 | NA | 1 | 2.76 |
| *Salmonella bongori* | 1 | 1.81 | 1 | 1.81 |
| *Salmonella enterica* | 3 | 3.59 | 2 | 3.63 |
| *Shigella dysenteriae* | 1 | 6.19 | 0 | NA |
| *Shigella flexneri* | 0 | NA | 1 | 6.51 |
| *Staphylococcus aureus* | 2 | 6.39 | 4 | 6.20 |
| *Streptococcus pneumoniae* | 1 | 7.26 | 1 | 7.28 |
| *Vibrio cholerae* | 7 | 5.66 | 8 | 6.03 |

<sup>1</sup>: *Campylobacter jejuni* or *Campylobacter coli*
<sup>2</sup>: *Shigella dysenteriae*, *Shigella boydii*, or *Shigella flexneri*

**Table S8:** Presence (1) and absence (0) of antibiotic resistance gene targets in Boston and Vellore by TAC. Ww=wastewater. MLS=macrolide, lincosamides, streptogramines. MRSA=methicillin-resistant Staphylococcus aureus.

| Target | Drug Class | Boston |  |  |  |  |  | Vellore |  |  |  |  |  |
| --- | --- | --- | --- | --- | --- | --- | --- | --- | --- | --- | --- | --- | --- |
|  |  | Ww 1 | Ww 2 | Ww 3 | Surface 1 | Surface 2 | Hospital | Ww 1 | Ww 2 | Surface 1 | Surface 2 | Surface 3 | Hospital |
| AAC(6)-Ib-cr | Aminoglycoside | 1 | 1 | 1 | 1 | 1 | 1 | 1 | 1 | 1 | 1 | 1 | 1 |
| aacC1 | Aminoglycoside | 1 | 1 | 1 | 0 | 1 | 1 | 1 | 1 | 1 | 1 | 1 | 1 |
| aacC2 | Aminoglycoside | 1 | 1 | 1 | 0 | 1 | 1 | 1 | 1 | 1 | 1 | 1 | 1 |
| aacC4 | Aminoglycoside | 1 | 1 | 1 | 0 | 0 | 1 | 0 | 1 | 1 | 1 | 1 | 1 |
| aphA6 | Aminoglycoside | 1 | 1 | 1 | 0 | 1 | 0 | 1 | 1 | 1 | 1 | 1 | 1 |
| ACC-1 group | Beta-lactam | 1 | 1 | 1 | 0 | 1 | 0 | 0 | 0 | 0 | 0 | 0 | 0 |
| ACC-3 | Beta-lactam | 1 | 1 | 1 | 0 | 1 | 0 | 0 | 0 | 0 | 1 | 0 | 0 |
| ACT 5/7 group | Beta-lactam | 1 | 1 | 1 | 0 | 1 | 1 | 1 | 1 | 1 | 1 | 1 | 1 |
| ACT-1 group | Beta-lactam | 1 | 1 | 1 | 0 | 1 | 1 | 1 | 1 | 1 | 1 | 1 | 1 |
| BES-1 | Beta-lactam | 1 | 1 | 1 | 0 | 1 | 1 | 1 | 1 | 1 | 1 | 1 | 1 |
| BIC-1 | Beta-lactam | 0 | 0 | 1 | 0 | 0 | 0 | 0 | 0 | 0 | 0 | 0 | 0 |
| ccrA | Beta-lactam | 1 | 1 | 1 | 0 | 1 | 0 | 1 | 1 | 1 | 1 | 1 | 1 |
| CFE-1 | Beta-lactam | 1 | 1 | 1 | 0 | 0 | 0 | 0 | 1 | 1 | 1 | 0 | 0 |
| CMY-10 Group | Beta-lactam | 1 | 1 | 1 | 0 | 1 | 1 | 1 | 1 | 1 | 1 | 1 | 1 |
| CTX-M-1 Group | Beta-lactam | 1 | 1 | 1 | 0 | 1 | 1 | 1 | 1 | 1 | 1 | 1 | 1 |
| CTX-M-8 Group | Beta-lactam | 1 | 0 | 1 | 0 | 0 | 0 | 0 | 1 | 1 | 1 | 1 | 1 |
| CTX-M-9 Group | Beta-lactam | 1 | 1 | 1 | 0 | 0 | 1 | 1 | 1 | 1 | 1 | 1 | 1 |
| DHA | Beta-lactam | 1 | 1 | 1 | 0 | 0 | 1 | 1 | 1 | 1 | 1 | 1 | 1 |
| FOX | Beta-lactam | 1 | 1 | 1 | 0 | 1 | 1 | 1 | 1 | 1 | 1 | 1 | 1 |
| GES | Beta-lactam | 1 | 1 | 1 | 1 | 1 | 1 | 1 | 1 | 1 | 1 | 1 | 1 |
| IMI & NMC-A | Beta-lactam | 0 | 0 | 0 | 0 | 0 | 0 | 0 | 0 | 1 | 0 | 0 | 0 |
| IMP-1 group | Beta-lactam | 0 | 0 | 0 | 0 | 0 | 1 | 1 | 1 | 1 | 1 | 1 | 1 |
| IMP-12 group | Beta-lactam | 0 | 0 | 0 | 1 | 1 | 1 | 1 | 1 | 1 | 0 | 1 | 1 |
| IMP-2 group | Beta-lactam | 0 | 1 | 1 | 1 | 1 | 0 | 1 | 0 | 1 | 0 | 0 | 1 |
| IMP-5 group | Beta-lactam | 1 | 0 | 1 | 1 | 1 | 1 | 1 | 1 | 1 | 0 | 1 | 1 |
| KPC | Beta-lactam | 1 | 1 | 1 | 1 | 1 | 1 | 1 | 1 | 1 | 1 | 1 | 1 |
| LAT | Beta-lactam | 1 | 1 | 1 | 0 | 1 | 1 | 1 | 1 | 1 | 1 | 1 | 1 |
| MIR | Beta-lactam | 1 | 1 | 1 | 0 | 1 | 1 | 1 | 1 | 1 | 1 | 1 | 1 |
| MOX | Beta-lactam | 1 | 1 | 1 | 1 | 1 | 1 | 1 | 1 | 1 | 1 | 1 | 1 |
| NDM | Beta-lactam | 0 | 0 | 0 | 0 | 0 | 0 | 1 | 1 | 1 | 1 | 1 | 1 |
| OXA-10 Group | Beta-lactam | 1 | 1 | 1 | 1 | 1 | 1 | 1 | 1 | 1 | 1 | 1 | 1 |
| OXA-18 | Beta-lactam | 1 | 1 | 1 | 0 | 0 | 1 | 0 | 0 | 0 | 0 | 0 | 0 |
| OXA-2 Group | Beta-lactam | 1 | 1 | 1 | 1 | 1 | 1 | 1 | 1 | 1 | 1 | 1 | 1 |
| OXA-23 Group | Beta-lactam | 1 | 1 | 1 | 0 | 1 | 0 | 0 | 1 | 1 | 1 | 1 | 1 |
| OXA-24 Group | Beta-lactam | 1 | 1 | 1 | 1 | 1 | 1 | 1 | 1 | 1 | 1 | 0 | 1 |
| OXA-45 | Beta-lactam | 0 | 0 | 0 | 0 | 0 | 0 | 0 | 0 | 0 | 0 | 0 | 0 |
| OXA-48 Group | Beta-lactam | 1 | 1 | 1 | 0 | 1 | 1 | 1 | 1 | 1 | 1 | 1 | 1 |
| OXA-50 Group | Beta-lactam | 1 | 1 | 1 | 0 | 1 | 1 | 1 | 1 | 1 | 1 | 1 | 1 |
| OXA-51 Group | Beta-lactam | 1 | 1 | 1 | 0 | 0 | 0 | 1 | 1 | 1 | 1 | 1 | 1 |
| OXA-54 | Beta-lactam | 0 | 0 | 0 | 0 | 1 | 0 | 0 | 0 | 0 | 0 | 0 | 0 |
| OXA-55 | Beta-lactam | 0 | 0 | 0 | 0 | 0 | 0 | 0 | 0 | 0 | 0 | 0 | 0 |
| OXA-58 Group | Beta-lactam | 1 | 1 | 1 | 1 | 1 | 1 | 1 | 1 | 1 | 1 | 1 | 1 |
| OXA-60 | Beta-lactam | 0 | 0 | 0 | 0 | 1 | 0 | 0 | 0 | 0 | 0 | 0 | 0 |
| Per-1 group | Beta-lactam | 1 | 1 | 1 | 0 | 0 | 1 | 1 | 1 | 1 | 1 | 1 | 1 |
| Per-2 group | Beta-lactam | 1 | 1 | 1 | 0 | 1 | 0 | 0 | 0 | 0 | 0 | 0 | 0 |
| SFC-1 | Beta-lactam | 0 | 0 | 0 | 0 | 0 | 0 | 0 | 0 | 0 | 0 | 0 | 0 |
| SFO-1 | Beta-lactam | 1 | 1 | 1 | 0 | 0 | 0 | 1 | 0 | 1 | 0 | 0 | 1 |
| SHV | Beta-lactam | 1 | 1 | 1 | 0 | 1 | 1 | 1 | 1 | 1 | 1 | 1 | 1 |
| SHV(156D) | Beta-lactam | 1 | 1 | 1 | 0 | 0 | 1 | 1 | 1 | 1 | 1 | 1 | 1 |
| SHV(238G240E) | Beta-lactam | 1 | 1 | 1 | 1 | 1 | 0 | 1 | 1 | 1 | 1 | 1 | 1 |
| SHV(238S240E) | Beta-lactam | 1 | 1 | 1 | 0 | 1 | 1 | 1 | 1 | 1 | 1 | 1 | 1 |
| SHV(238S240K) | Beta-lactam | 1 | 1 | 1 | 0 | 1 | 1 | 1 | 1 | 1 | 1 | 1 | 1 |
| SME | Beta-lactam | 0 | 0 | 0 | 0 | 0 | 0 | 0 | 0 | 0 | 0 | 0 | 0 |
| TLA-1 | Beta-lactam | 1 | 1 | 1 | 0 | 1 | 1 | 1 | 1 | 1 | 1 | 1 | 1 |
| VIM-1 group | Beta-lactam | 1 | 1 | 1 | 0 | 1 | 1 | 1 | 1 | 1 | 1 | 1 | 1 |
| VIM-13 | Beta-lactam | 1 | 1 | 1 | 0 | 0 | 1 | 1 | 1 | 1 | 1 | 1 | 1 |
| VIM-7 | Beta-lactam | 0 | 0 | 0 | 0 | 0 | 0 | 0 | 0 | 0 | 0 | 0 | 0 |
| ereB | Erythromycin | 1 | 1 | 1 | 0 | 1 | 0 | 1 | 1 | 1 | 1 | 1 | 1 |
| QepA | Fluoroquinolone | 0 | 0 | 0 | 0 | 0 | 0 | 0 | 1 | 1 | 0 | 0 | 1 |
| QnrA | Fluoroquinolone | 1 | 1 | 1 | 1 | 1 | 1 | 1 | 1 | 1 | 1 | 1 | 1 |
| QnrB-1 group | Fluoroquinolone | 1 | 1 | 1 | 0 | 1 | 1 | 1 | 1 | 1 | 1 | 1 | 1 |
| QnrB-31 group | Fluoroquinolone | 0 | 0 | 0 | 0 | 0 | 0 | 0 | 0 | 0 | 0 | 0 | 0 |
| QnrB-4 group | Fluoroquinolone | 1 | 1 | 1 | 0 | 0 | 1 | 1 | 1 | 1 | 1 | 1 | 1 |
| QnrB-5 group | Fluoroquinolone | 1 | 1 | 1 | 0 | 1 | 1 | 0 | 1 | 1 | 1 | 1 | 1 |
| QnrB-8 group | Fluoroquinolone | 1 | 1 | 1 | 0 | 1 | 1 | 0 | 1 | 1 | 1 | 0 | 1 |
| QnrC | Fluoroquinolone | 0 | 0 | 0 | 0 | 0 | 0 | 0 | 0 | 0 | 0 | 0 | 0 |
| QnrD | Fluoroquinolone | 1 | 1 | 1 | 0 | 0 | 1 | 0 | 1 | 1 | 1 | 1 | 1 |
| QnrS | Fluoroquinolone | 1 | 1 | 1 | 1 | 1 | 1 | 1 | 1 | 1 | 1 | 1 | 1 |
| ermA | MLS | 1 | 1 | 1 | 0 | 1 | 0 | 0 | 1 | 1 | 1 | 1 | 1 |
| ermB | MLS | 1 | 1 | 1 | 1 | 1 | 1 | 1 | 1 | 1 | 1 | 1 | 1 |
| ermC | MLS | 1 | 1 | 1 | 0 | 0 | 1 | 0 | 1 | 0 | 1 | 0 | 1 |
| mefA | MLS | 1 | 1 | 1 | 1 | 1 | 1 | 1 | 1 | 1 | 1 | 1 | 1 |
| msrA | MLS | 1 | 1 | 1 | 1 | 1 | 1 | 1 | 0 | 0 | 1 | 0 | 1 |
| lukF | MRSA | 0 | 0 | 0 | 0 | 0 | 0 | 0 | 0 | 0 | 0 | 0 | 0 |
| mecA | MRSA | 0 | 1 | 0 | 1 | 1 | 1 | 0 | 1 | 0 | 1 | 1 | 1 |
| oprj | Multidrug efflux pump | 1 | 1 | 1 | 0 | 0 | 1 | 0 | 1 | 0 | 1 | 0 | 1 |
| oprM | Multidrug efflux pump | 1 | 1 | 1 | 1 | 1 | 1 | 0 | 1 | 1 | 1 | 1 | 1 |
| tetA | Tetracycline | 1 | 1 | 1 | 1 | 1 | 1 | 1 | 1 | 1 | 1 | 1 | 1 |
| tetB | Tetracycline | 1 | 1 | 1 | 0 | 1 | 1 | 0 | 1 | 1 | 1 | 1 | 1 |
| vanB | Vancomycin | 1 | 1 | 1 | 0 | 1 | 1 | 0 | 0 | 0 | 0 | 0 | 1 |
| vanC | Vancomycin | 0 | 1 | 1 | 0 | 0 | 0 | 0 | 1 | 1 | 1 | 0 | 1 |
| VEB | Vancomycin | 1 | 1 | 1 | 1 | 1 | 1 | 1 | 1 | 1 | 1 | 1 | 1 |

**Table S9:** Conditional probabilities comparing raw long-reads (unassembled and unpolished) to real-time PCR for antibiotic resistance gene identification.

| Gene | Sensitivity | Specificity | Positive Predictive Value | Negative Predictive Value |
| --- | --- | --- | --- | --- |
| CTX-M-1 | 0.00 | 100.00 | NA | 8.33 |
| CTX-M-8 | 0.00 | 100.00 | NA | 41.67 |
| CTX-M-9 | 0.00 | 100.00 | NA | 16.67 |
| KPC | 25.00 | NA | 100.00 | 0.00 |
| *mecA* | 0.00 | 100.00 | NA | 33.33 |
| NDM | 16.67 | 100.00 | 100.00 | 54.55 |
| OXA-10 | 58.33 | NA | 100.00 | 0.00 |
| OXA-18 | 0.00 | 100.00 | NA | 66.67 |
| OXA-2 | 50.00 | NA | 100.00 | 0.00 |
| OXA-23 | 0.00 | 100.00 | NA | 25.00 |
| OXA-24 | 18.18 | 100.00 | 100.00 | 10.00 |
| OXA-45 | NA | 100.00 | NA | 100.00 |
| OXA-48 | 9.09 | 100.00 | 100.00 | 9.09 |
| OXA-50 | 0.00 | 100.00 | NA | 8.33 |
| OXA-51 | 0.00 | 100.00 | NA | 25.00 |
| OXA-54 | 0.00 | 100.00 | NA | 91.67 |
| OXA-55 | NA | 100.00 | NA | 100.00 |
| OXA-58 | 66.67 | NA | 100.00 | 0.00 |
| OXA-60 | 0.00 | 100.00 | NA | 91.67 |
| *qnrS* | 33.33 | NA | 100.00 | 0.00 |
| SHV-1 | 0.00 | 100.00 | NA | 8.33 |
| blaSHV-27 | 0.00 | 100.00 | NA | 16.67 |
| blaSHV-31 | 0.00 | 100.00 | NA | 58.33 |
| blaSHV-2 | 0.00 | 100.00 | NA | 8.33 |
| blaSHV-12 | 9.09 | 100.00 | 100.00 | 9.09 |
| *tetA* | 66.67 | NA | 100.00 | 0.00 |
| *tetB* | 50.00 | 100.00 | 100.00 | 28.57 |
| *vanC* | 0.00 | 100.00 | NA | 50.00 |

**Table S10:** Concordance between ARG identification on polished and unpolished contigs at the allele and gene level by drug class. MLS=macrolide, lincosamides, streptogramines.

| Drug Class | Concordance (Allele) | Concordance (Gene) | Total Hits |
| --- | --- | --- | --- |
| Beta-lactamase | 0.78 | 0.96 | 178 |
| Aminoglycoside | 0.88 | 0.95 | 168 |
| Folate pathway antagonist | 0.84 | 1.00 | 56 |
| Macrolide | 0.95 | 0.99 | 134 |
| MLS | 0.92 | 1.00 | 38 |
| Phenicol | 0.88 | 1.00 | 25 |
| Polymyxin | 0.67 | 1.00 | 15 |
| Quinolone | 0.89 | 0.95 | 37 |
| Tetracycline | 0.90 | 0.98 | 111 |

